# Predicting Prognosis in COVID-19 Patients using Machine Learning and Readily Available Clinical Data

**DOI:** 10.1101/2021.01.29.21250762

**Authors:** Thomas W. Campbell, Melissa P. Wilson, Heinrich Roder, Samantha MaWhinney, Robert W. Georgantas, Laura K. Maguire, Joanna Roder, Kristine M. Erlandson

**Author notes:** Corresponding Author. Address: 2970 Wilderness Place, Suite 100, Boulder, CO 80301. co-senior authors (Joanna Roder, Kristine M. Erlandson). Contributions: conceptualization, formal analysis, methodology, software, writing – original draft, writing – review and editing. Contributions: Data extraction, data harmonization, evaluation of outcome variable extraction. Contributions: supervision, methodology, software, writing – original draft, writing – review and editing. Contributions: cohort development, statistical and analysis review, manuscript review and editing. Contributions: supervision, writing – review and editing. Contributions: formal analysis, writing – review and editing. Contributions: supervision, methodology, formal analysis, writing – original draft, writing – review and editing. Contributions: cohort development, data collection, conceptualization of project, review and editing of initial and subsequent manuscript drafts.

## Abstract

**Rationale:** Prognostic tools for aiding in the treatment of hospitalized COVID-19 patients could help improve outcome by identifying patients at higher or lower risk of severe disease.

**Objectives:** The study objective was to develop models to stratify patients by risk of severe outcomes during COVID-19 hospitalization using readily available information at hospital admission.

**Methods:** Hierarchical ensemble classification models were trained on a set of 229 patients hospitalized with COVID-19 to predict severe outcomes, including ICU admission, development of ARDS, or intubation, using easily attainable attributes including basic patient characteristics, vital signs at admission, and basic lab results collected at time of presentation. Each test stratifies patients into groups of increasing risk. An additional cohort of 330 patients was used for blinded, independent validation. Shapley value analysis evaluated which attributes contributed most to the models’ predictions of risk.

**Measurements and Main Results:** Test performance was assessed using precision (positive predictive value) and recall (sensitivity) of the final risk groups. All test cut-offs were fixed prior to blinded validation. In both development and validation, the tests achieved precision in the lowest risk groups near or above 0.9. The proportion of patients with severe outcomes significantly increased across increasing risk groups. While the importance of attributes varied by test and patient, CRP, LDH, and D-dimer were often found to be important in the assignment of risk label.

**Conclusions:** Risk of severe outcomes for patients hospitalized with COVID-19 infection can be assessed using machine learning-based models based on attributes routinely collected at hospital admission.

## Introduction

The coronavirus disease (COVID-19) pandemic continues to place world-wide health systems under pressure, with COVID-19 hospitalization rates in the USA exceeding 240 per 100,000 population (1). Better understanding of how to identify patients at greatest risk of developing severe lung disease, requiring intubation, or dying from COVID-19 would help inform discussions of the risks and benefits of treatments (bamlanivmad (2), remdesivir (3), others (4)) with patients and family members, help target higher risk populations for clinical trials of therapeutic agents, compare outcomes between patients with similar risk, and prioritize resources in a time of shortage.

As providers have gained experience treating patients hospitalized with COVID-19, multiple factors have been identified as associated with risk of hospitalization, severe disease, or mortality, including older age and certain comorbidities (5-7). Laboratory results, such as neutrophil-to-lymphocyte ratio (NLR), d-dimer and C-reactive protein (CRP) (5,8-9) have been associated with poor outcomes. However, as the COVID-19 patient profile has changed during the pandemic, the relative influence of individual risk factors has been difficult to elucidate.

While traditional statistical modelling has been used to combine multiple attributes to predict outcomes for patients with COVID-19 (10-11), modern machine learning (ML) has added advantage in its capability to discover more complex interactions between correlated attributes, giving ML greater power to discriminate outcomes. Such methods have shown promise for predicting acute respiratory distress syndrome (ARDS) in diseases other than COVID-19 (12-13). Multiple studies have demonstrated the potential utility of ML for predicting poor outcomes in COVID-19. An early systematic review (14) noted that comorbidities, gender, CRP, and creatinine frequently appeared as attributes selected during prognostic ML-based modelling. More recently published studies confirm the potential utility of complex ML-based tests to predict severe COVID-19 or mortality (15-20).

Many patients hospitalized with COVID-19 provide limited medical history due to not engaging in routine care, not having easily accessible comorbidities data, or being intubated shortly after arrival. Many of the complex ML models may not be able to assess a large proportion of patients due to missing data. Hence, the most useful test to assess risk of complications and severe disease at the point of hospital admission would require only attributes that can be easily and quickly collected at the time of admission.

The goal of this study was to create and validate tests to risk stratify COVID-19 patients for progression to severe disease using only readily available clinical, demographic, and laboratory attributes collected at the point of hospital admission. Given the nature of these attributes, the tests stratifying risk of admission to the intensive care unit (ICU), intubation, or progression to ARDS could be validated, blinded to outcome, on an independent patient cohort using data drawn from electronic health records (EHRs).

## Methods

### Data Extraction and Patient Cohorts

For test development, data was collected for all patients 18 years or older admitted to the University of Colorado Health (UCH) hospital in Aurora, CO with a positive COVID-19 nasopharyngeal polymerase chain reaction test between mid-March and mid-May 2020. Patient information for attributes deemed potentially useful at the time was extracted from the EHR by medical students and stored in a REDCap database (21). Patient data was restricted to first recorded admission, and to first recorded observations for laboratory values and vital measures.

The deidentified dataset was transferred to Biodesix for test development. A set of 26 attributes (indicated with * in Table 1) to be used in classifier training was selected, limited to attributes routinely collected at hospital admission and available in the EHR, where inclusion of an attribute did not reduce the size of the cohort with complete data by more than 10%. The development cohort (Figure 1, Table E1) included patients with complete information for the selected attributes.

**Table 1:** Patient characteristics for the development and independent validation cohorts.

|  |  | Development Cohort<br>(N=229) | Validation Cohort<br>(N=330) |
| --- | --- | --- | --- |
| Categorical Attribute | Class | n (%) | n (%) |
| Race* | White | 41 (17.9) | 113 (34.2) |
|  | Black/African American | 52 (22.7) | 23 (7.0) |
|  | Hispanic/Latino | 94 (41.0) | 164 (49.7) |
|  | Other | 31 (13.5) | 25 (7.6) |
|  | Unknown | 11 (4.8) | 5 (1.5) |
| Sex* | Male | 124 (54.1) | 172 (52.1) |
|  | Female | 105 (45.9) | 158 (47.9) |
| eGFR* | $\geq 60$ mL/min/1.73m<sup>2</sup> | 180 (78.6) | 244 (73.9) |
| | $30 \geq$ but $<60$ mL/min/1.73m<sup>2</sup> | 34 (14.8) | 61 (18.5) |
| | $< 30$ mL/min/1.73m<sup>2</sup> | 15 (6.6) | 25 (7.6) |
| Hypertension | Yes | 100 (43.7) | NA |
|  | No | 126 (56.3) | NA |
| Diabetes | Yes | 82 (35.8) | NA |

Machine Learning COVID-19 Risk Stratification
|  | No | 147 (64.2) | NA |
| --- | --- | --- | --- |
| **Continuous Attribute** |  | **median (Q1-Q3)** | **median (Q1-Q3)** |
| BMI**, kg/m<sup>2</sup> |  | 30 (27-36) |  |
| Age*, years |  | 57 (43-68) | 57 (44-70) |
| Temperature*, °C |  | 37 (37-38) | 37 (37-38) |
| Heart Rate*,<br>beats/minute |  | 98 (84-110) | 98 (85-110) |
| Systolic *, mm Hg |  | 130 (120-150) | 130 (120-140) |
| Diastolic BP*, mm Hg |  | 74 (65-83) | 74 (65-84) |
| Respiratory Rate*,<br>breaths/minute |  | 20 (18-24) | 20 (18-24) |
| Oxygen Saturation*, % |  | 92 (87-94) | 92 (88-95) |
| Weight*, kg |  | 82 (72-100) | 85 (73-100) |
| QTc*, |  | 440 (420-460) | 440 (420-470) |
| Sodium*, mmol/L |  | 140 (130-140) | 140 (130-140) |
| Potassium*, mmol/L |  | 3.8 (3.4-4.0) | 3.9 (3.6-4.2) |
| Carbon dioxide* mmol/L |  | 23 (21-25) | 23 (21-25) |
| BUN* mg/dL |  | 13 (10-20) | 15 (10-22) |
| Creatinine* mg/dL |  | 0.94 (0.69-1.2) | 0.87 (0.72-1.2) |
| Anion Gap*, mmol/L |  | 12 (10-13) | 11 (9-13) |
| WBC Count* x 10<sup>9</sup> cells/L |  | 6.8 (5.3-8.9) | 7 (5.5-9.1) |
| Hemoglobin* g/dL |  | 15 (13-16) | 14 (13-15) |
| Hematocrit*, % |  | 44 (40-47) | 42 (38-46) |
| Platelet Count* x10<sup>9</sup><br>cells/L |  | 210 (160-260) | 200 (160-260) |
| LDH* U/L |  | 320 (260-420) | 450 (290-750) |
| D-Dimer* ng/mL |  | 860 (530-1500) | 740 (410-1500) |
| CRP*, mg/L |  | 83 (41-150) | 79 (37-160) |
| Ferritin*, ng/mL |  | 360 (170-730) | 310 (130-600) |

Definition of abbreviations: eGFR = estimated glomerular filtration rate; BP = blood pressure; QTc = corrected QT interval; BUN = blood urea nitrogen; CO<sub>2</sub>= carbon dioxide; WBC = white blood cell; LDH = lactate dehydrogenase; CRP = C-reactive protein; BMI = body mass index
\*Used for classification
\*\* only complete for 205 out 229 patients

**Figure 1.**
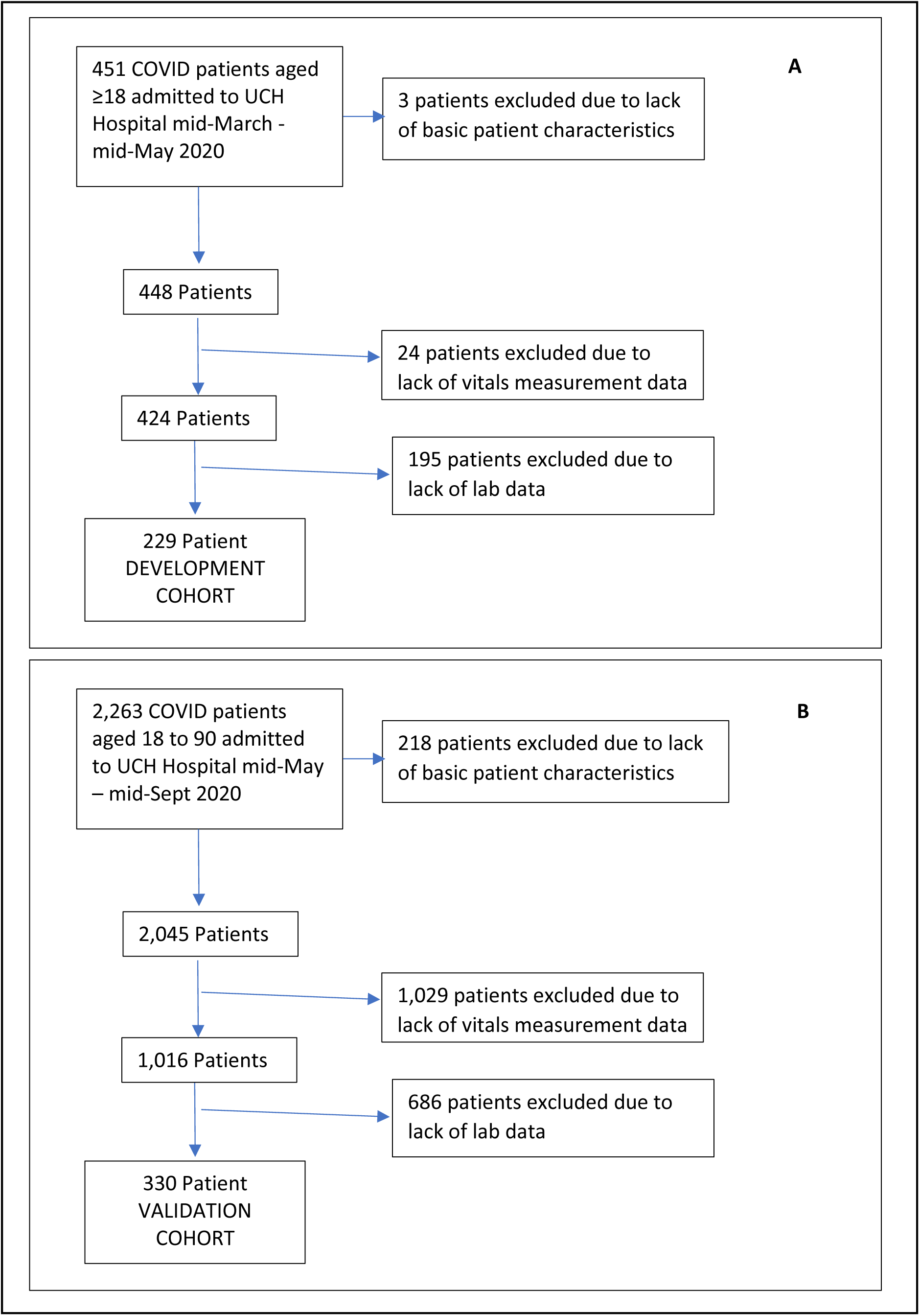
consort diagrams of patient selection down to development (A) and validation (B) cohorts.

An independent validation cohort was derived from data provided by UCH’s EHR data warehouse service. The validation cohort included all patients aged between 18 and 90 years, hospitalized in a UCH system facility with a positive COVID-19 test or diagnosis, with first admission between mid-May 2020 and mid-September 2020 with complete data on the 26 required attributes (Figure 1). Deidentified data for the validation cohort were transferred to Biodesix for blinded test classification generation. Outcome data for test performance evaluation for the validation cohort was only shared after test classifications was returned to UCH investigators. More details on data curation and validation are provided in the supplement. The study was reviewed and approved by the Colorado Multiple Institutional Review Board.

### Test Development

For each endpoint considered, a series of classifiers were trained to classify patients as higher or lower risk for the endpoint (ICU admission, intubation, ARDS) using the same set of 26 attributes. These classifiers were arranged in a hierarchy (Figure 2) to give a final classification of up to four possible risk groups. The fundamental model behind each component classifier was the Diagnostic Cortex^®^ platform, a strongly dropout-regularized, feature abstracted, ensemble logistic regression (22). The classifier at the top of each test’s hierarchy was augmented with an additional decision tree model. Reliable results were obtained in the development cohort using out-of-bag estimates (23). ML methods are described fully in the supplement (fig E1-E3).

**Figure 2:**
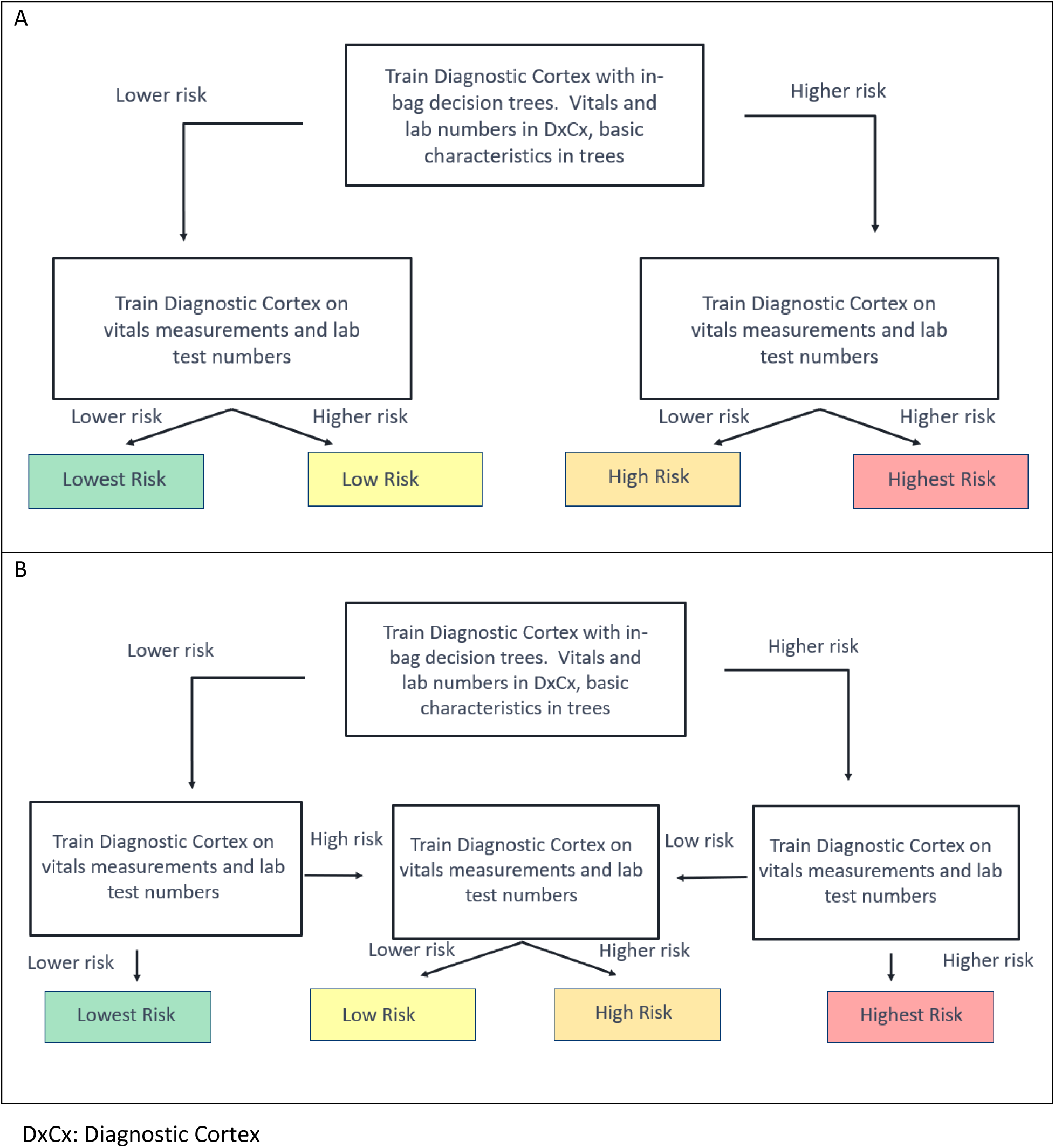
Hierarchical Configuration of Classifiers used for Risk Assessment for Each Endpoint. A Diagnostic Cortex model with in-bag decision tree model (represented by the top box) was used to stratify the entire development cohort into a higher and lower risk group for each endpoint. Diagnostic Cortex models (middle boxes) without trees were used to split the resulting two groups further according to one of the two schemas. (Schema A was used for the tests predicting risk of any complication and intubation. Schema B was used for the tests predicting risk of ARDS and admission to the ICU.)

The relative importance of the attributes used in the classification generated for an individual patient was evaluated using Shapley Values (24, 25) for 50 patients in the validation cohort chosen to span different test classification groups (figures E18-E25).

### Statistical Methods

The validation cohort was analyzed following a prespecified statistical analysis plan, included in the supplement. Statistical analyses were performed using SAS Enterprise Guide 8.2 (SAS 9.4) (SAS Institute, Cary, NC). Cochran-Armitage test and Fisher’s exact test were used to assess trends and differences in proportions. Confidence intervals (CIs) for proportions were calculated using the Clopper-Pearson method.

## Results

Patients included in the development cohort were generally similar to those excluded due to lack of complete data, although rate of intubation and systolic blood pressure were slightly higher and oxygen saturation and d-dimer level lower. Patients included in the validation cohort exhibited higher rates of severe outcomes than patients excluded due to lack of complete data and multiple laboratory measurements and vital signs indicated inferior prognosis (tables E2-E5).

Two-hundred twenty-nine patients were included in the development cohort: 77 (34%) were admitted to the ICU, 53 (23%) were intubated, and 45 (20%) developed ARDS. The proportions of patients experiencing poor outcomes were smaller in the 330 patients in the validation cohort, of whom 85 (26%) were admitted to the ICU, 42 (13%) were intubated, and 35 (11%) developed ARDS. The patient characteristics of the two cohorts are summarized in Table 1. All attributes used in classification are contained in Table 1 and denoted with an asterisk. The time from data collection to ICU admission was estimated in the validation cohort and is summarized in figure 3.

**Figure 3:**
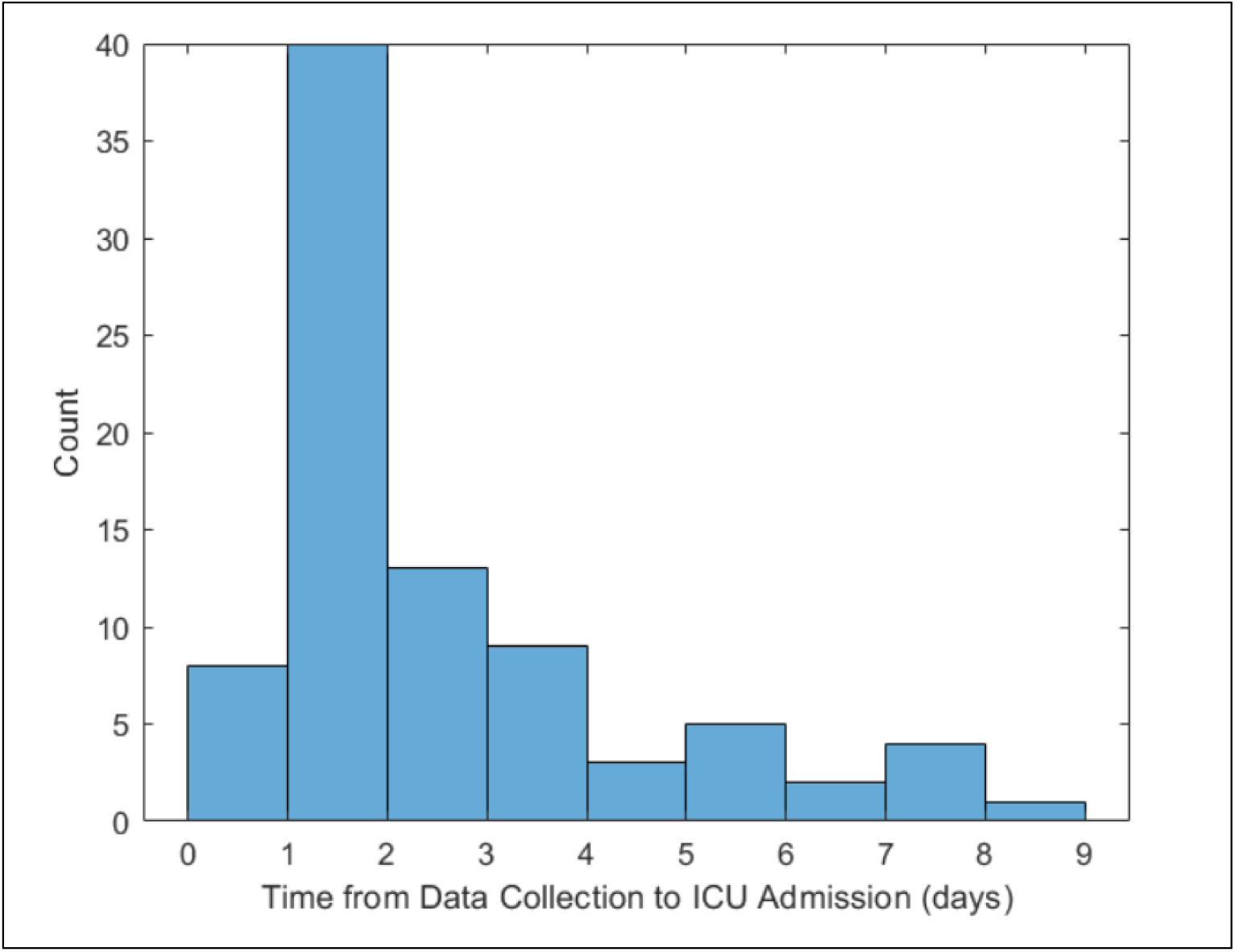
Time from data collection to admission to the ICU for the 85 patients admitted to the ICU in the validation cohort. A time in the [0,1] bin indicates the patient was admitted on the same day as the data was collected.

Males were slightly overrepresented in both cohorts (54% in development and 52% in validation). The median age in both cohorts was 57 years. The racial/ethnic composure of both cohorts were skewed compared with the overall Colorado demographics (26) with White being significantly underrepresented in both sets, and Black being overrepresented only in the development cohort.

### Predicting ICU Admission

Test development for prediction of ICU admission used the schema in Figure 2B, with the flow of patients through the hierarchical classification scheme shown in Figure 4. The test classified 73 (32%), 54 (24%), 51 (22%), and 51 (22%) of the development cohort to the lowest, low, high, and highest risk groups, respectively. The proportion of patients admitted to the ICU increased with increasing risk group. Only 12% of patients in the lowest risk group were admitted to the ICU, whereas 65% of the highest risk group were admitted. Performance was similar in the validation cohort, of which 28%, 22%, 27%, and 22% were classified to the lowest, low, high, and highest risk groups, respectively. The proportions of patients that were admitted to the ICU were significantly associated with increasing risk subgroup (Cochran-Armitage p < 0.0001). The proportion of patients admitted to the ICU in the lowest risk group was 11% (95% CI: 5%-19%), significantly lower than that in the other groups (Fisher’s exact p < 0.0001). The proportion of patients admitted to the ICU in the highest risk group was 51% (95% CI: 39%-63%), Fisher’s exact p (highest risk vs other) < 0.0001).

**Figure 4:**
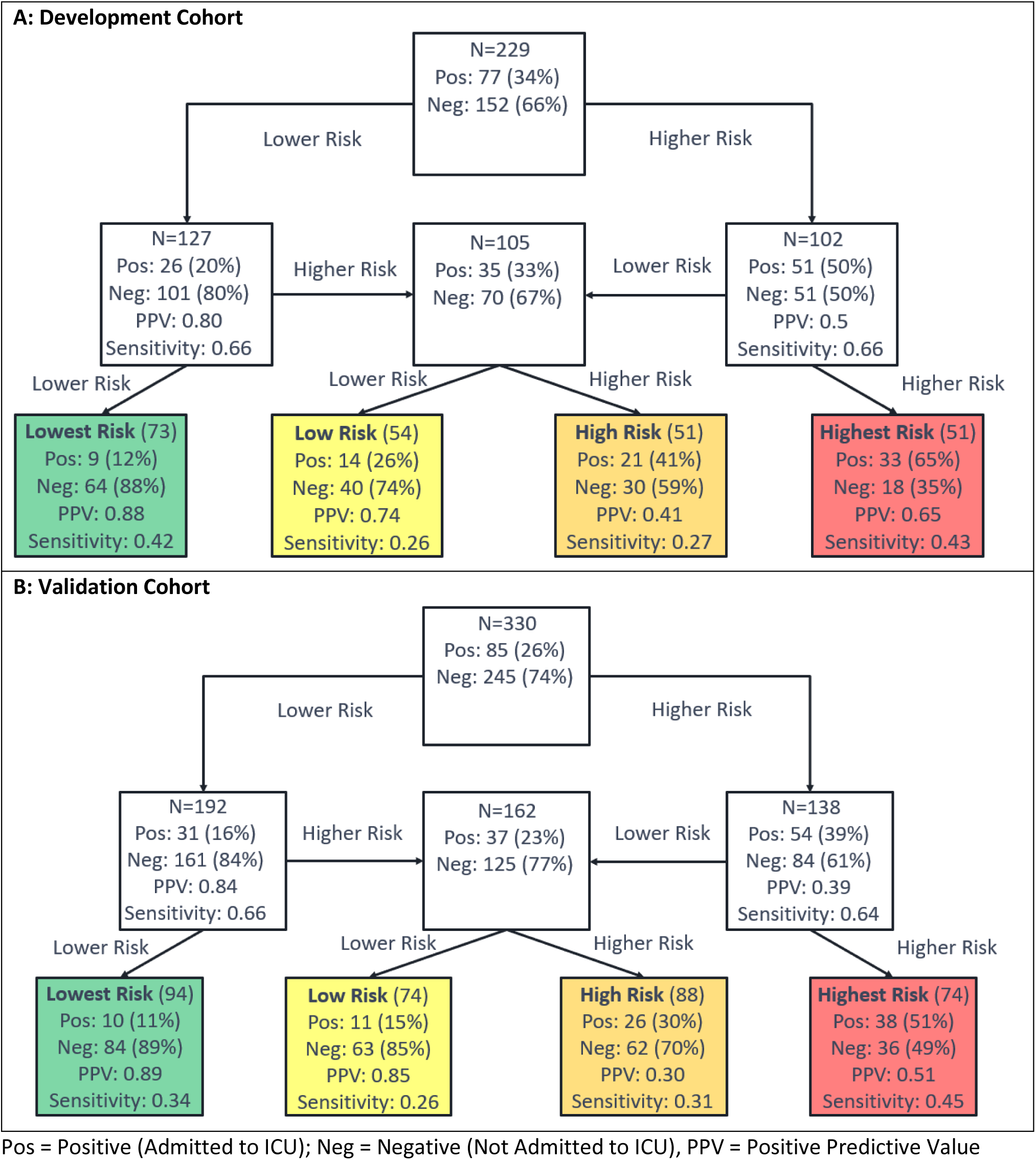
Performance Flow Chart for the Risk Assessment Test for ICU Admission for (A) the Development Cohort and (B) the Validation Cohort. Each uncolored box represents a classifier with the contents reflecting the set of patients to be classified by the classifier. The colored boxes represent the final risk groups with the contents reflecting composition of the groups and test performance.

Test classification was associated with heart rate, respiratory rate, oxygen saturation, sodium, blood urea nitrogen (BUN), anion gap, white blood cell (WBC) count, lactate dehydrogenase (LDH), d-dimer, CRP, and ferritin (tables E6-E9). Shapley Value analysis (figure E19) showed that the attributes most important for classification generation varied by patient, but that lowest risk classification was most often explained by CRP and d-dimer, with ferritin, LDH, platelet count, heart rate, respiratory rate, oxygen saturation, and WBC also of relatively higher importance. In highest risk patients, classification was related to LDH and d-dimer, although oxygen saturation, respiratory rate, CRP, ferritin, and weight were also important for some patients.

### Predicting ARDS

Test development for prediction of ARDS used the schema of Figure 2B, with the flow of patients through the hierarchical classification scheme shown in Figure 5. Note that, for this test, the lowest and lower risk groups were combined into a single lowest risk group, as the proportion of patients with ARDS was found to be similar in both subgroups during test development. The test predicting risk of developing ARDS classified 142 (62%), 47 (21%), and 40 (17%) patients to the lowest, low, and highest risk groups, respectively, in the development cohort. The proportion of patients developing ARDS increased across risk groups. In the lowest risk group, only 8% of patients developed ARDS, while in the highest risk group 45% of patients developed ARDS. The proportions of patients assigned to the lowest, high, and highest risk groups were similar in the validation cohort: 190 (58%), 73 (22%), and 67 (20%), respectively. The proportion of patients that developed ARDS increased with increasing risk group (Cochran-Armitage p = 0.02). Although the lowest risk group contained more than half of the patients, only 5% (95% CI: 3%-9%) of them developed ARDS (Fisher’s exact p (lowest vs other) < 0.0001). In contrast, the percentage of patients developing ARDS in the highest risk group was 27% (95% CI: 17%-39%), Fisher’s exact p (highest vs other) = 0.0004.

**Figure 5:**
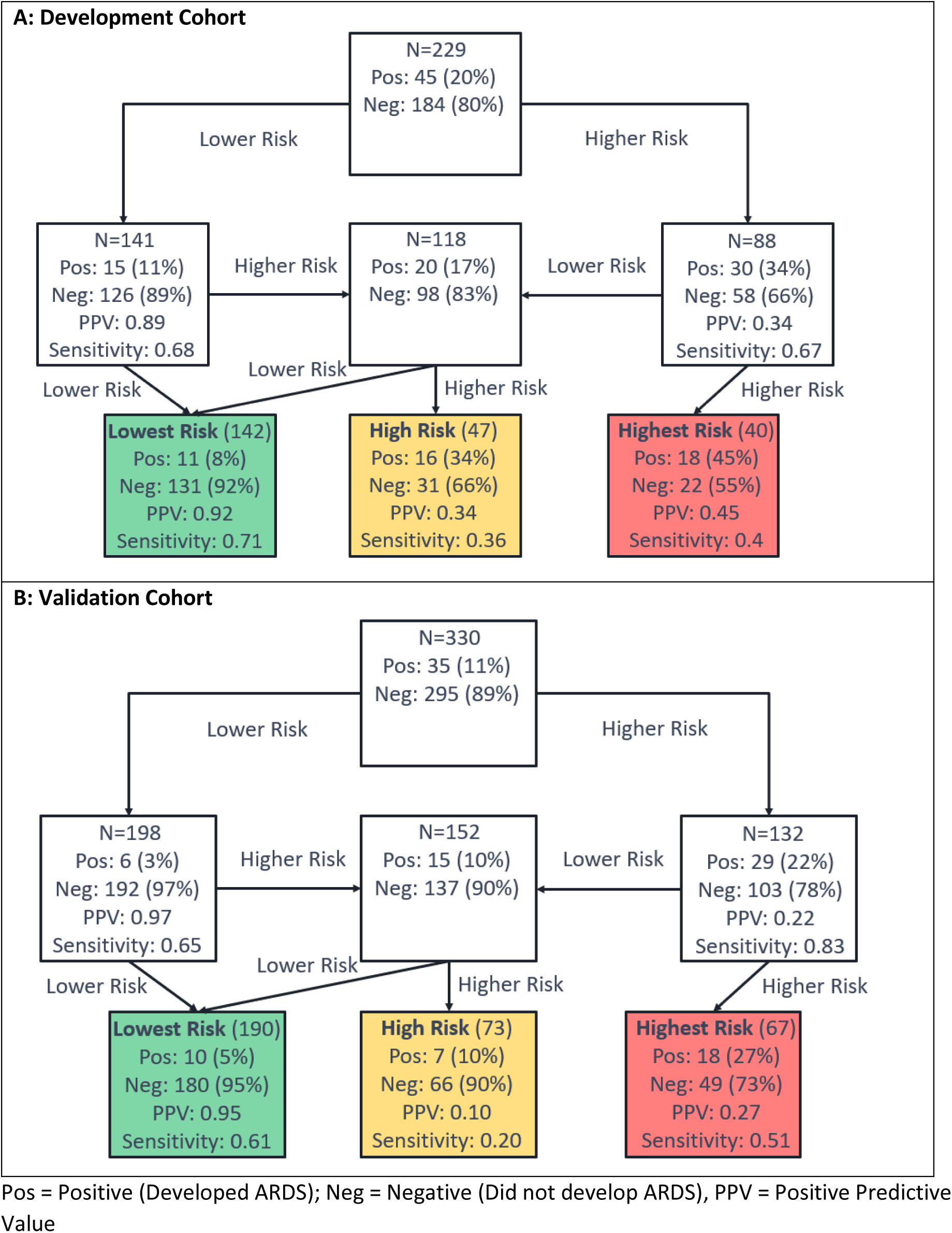
Performance Flow Chart for the Test Predicting Risk of Developing ARDS in (A) the Development Cohort, (B) the Validation Cohort. Each uncolored box represents a classifier with the contents reflecting the set of patients to be classified by the classifier. The colored boxes represent the final risk groups with the contents reflecting composition of the groups and test performance.

Test classification was associated with age, blood pressure, oxygen saturation, sodium, BUN, WBC, LDH, d-dimer, CRP, and ferritin (tables E10-E13). Shapley value analysis (figure E18) again showed that the attributes most important for classification generation varied by patient, but that CRP and d-dimer contributed substantially to a classification of lowest risk, with race and age also contributing for some patients. Highest risk classification was related to LDH, CRP, and BUN in most patients and oxygen saturation in some patients.

### Predicting Intubation

Test development to predict intubation used the schema of Figure 2A, with the flow of patients through the hierarchical classification scheme shown in Figure 6. The test predicting risk of intubation classified 74 (32%), 62 (27%), 55 (24%), and 38 (17%) patients of the development cohort to the lowest, low, intermediate, and highest risk groups, respectively and 86 (26%), 120 (36%), 67 (20%), and 57 (17%), respectively, in the validation cohort. The percentage of patients that were intubated increased from 9% to 16% to 24% to 61% across increasing risk groups. The patients of the validation cohort were assigned 86 (26%):120 (36%):67 (20%): 57 (17%) to the lowest, low, high and highest risk groups. Only 1% (95% CI: 0%-6%) of patients in the lowest risk group were intubated (Fisher’s exact p (vs other <0.0001), while 33% (95%CI: 21%-47%) of patients in the highest risk group were intubated (Fisher’s exact p (vs other) <0.0001). There was a significant association of increase in intubation with increasing risk group (Cochran-Armitage p <0.0001).

**Figure 6:**
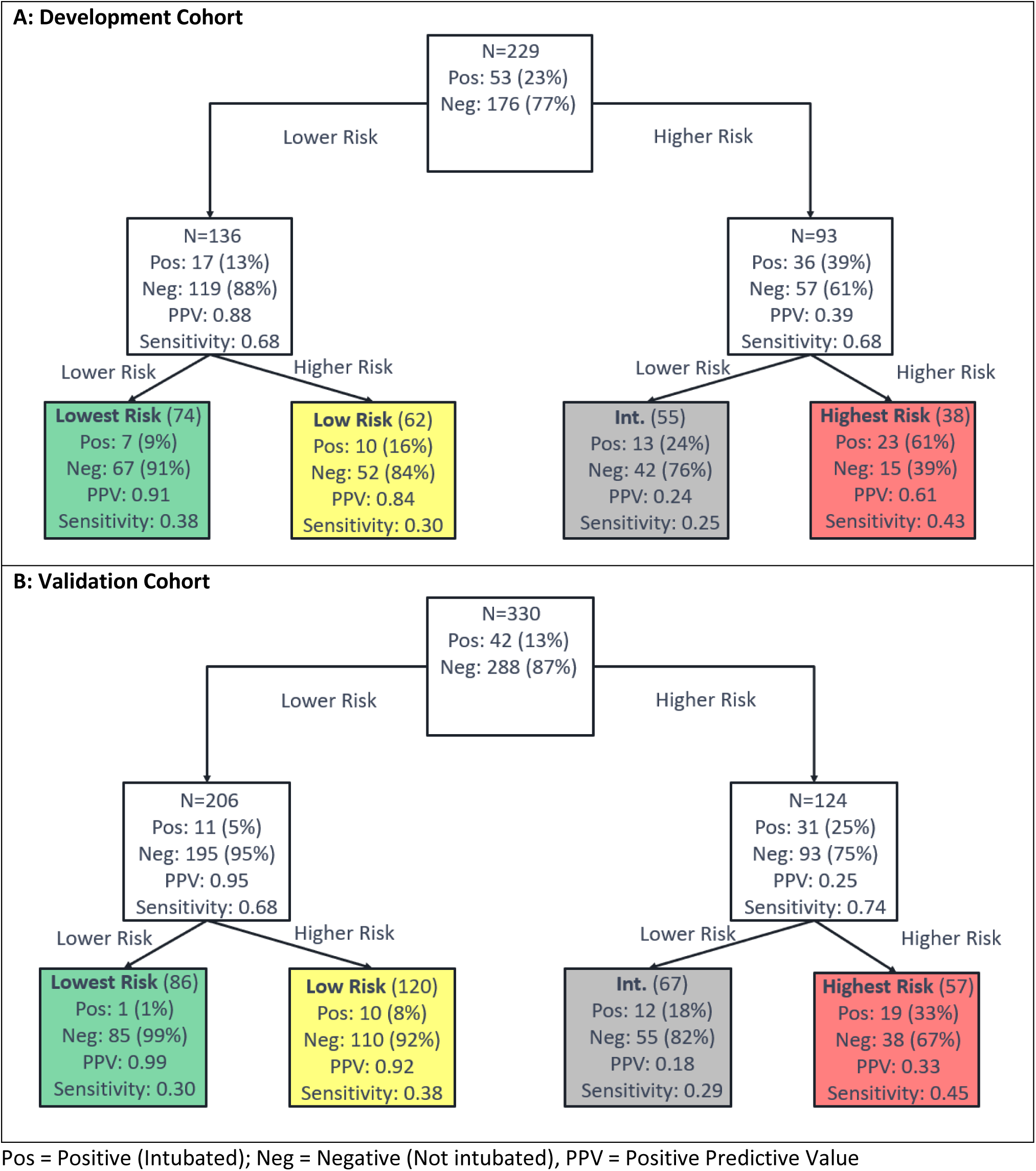
Performance Flow Chart for the Test Assessing Risk of Intubation for (A) the Development Cohort and (B) the Validation Cohort. Each uncolored box represents a classifier with the contents reflecting the set of patients to be classified by the classifier. The colored boxes represent the final risk groups with the contents reflecting composition of the groups and test performance.

Test classification was associated with respiratory rate, oxygen saturation, BUN, creatinine, anion gap, WBC, LDH, D-dimer CRP, and ferritin (tables E14-17). Shapley value analysis (figure E20) revealed that multiple attributes, varying by patient, were of increased relevance for classification. However, CRP was found to be generally most relevant for generation of a lowest risk classification, with d-dimer, LDH, BUN, race, and age also important factors. A highest risk classification was related to LDH, oxygen saturation, CRP, and BUN.

### Predicting Any Complication

Test development to predict the risk of any complication during hospitalization used the schema of Figure 2A. Performance in development, shown in detail in figures E15-E17 and tables E18-E19, was similar to that of the other tests presented. We did not seek to validate this test on the validation cohort, as accurate extraction of this endpoint from the EHRs alone was not possible.

### Predicting Mortality

Mortality was scarce in the development cohort, with only 23 deaths out of 220 patients with complete data. For this reason, mortality was not considered a viable endpoint for classifier training. The performance of the classifiers developed for other endpoints in predicting mortality was evaluated. All four tests performed similarly, see supplement, with the test predicting risk of intubation having the best performance. Only one out of 70 patients in the lowest risk group (1%) died, and 12 out of 37 patients in the highest risk group (32%) died.

### Comparison with other tests

An attempt was made to compare the performance of the tests to similar published classifiers (9, 11, 16). The classifier described by Wu et al. (16) was most directly comparable, but we were unable to complete the comparison due to lack of required attributes. The classifier described in Xiao et al. (11) was a binary classifier predicting lower or higher risk of severe disease. This classifier applied on the development cohort yielded very similar areas under the receiver-operating characteristic (ROC) curves to those for the first classifiers in our hierarchies for each endpoint. Using the specified cutoff (11), relatively small, but pure lowest risk groups were observed for all four endpoints. While the purity was similar to that of our tests, the lowest risk group sizes were roughly half the size. The classifier described by Liu et al. (9) was simple, using only age and NLR to stratify patients into four risk groups. Across all four endpoints, purities in the lowest risk groups were similar to those presented here but were roughly half the size. The highest risk groups were nearly twice the size of those for our tests but had purities a factor of 2-3 smaller. Performance for predicting ICU admission was poor, with the low risk group containing a similar proportion of patients admitted to the ICU as the highest risk group.

## Discussion

Using readily accessible EHR-derived data recorded at the time of hospital admission, we developed and blindly validated three different ML-based risk tests predicting three measures of progression to severe disease in patients hospitalized with COVID-19; namely risk of progression to ICU admission, intubation, and ARDS diagnosis.

Multiple previous studies have used ML-based algorithms to predict COVID-19 prognosis by combining basic patient demographics, vital signs, laboratory measurements, and comorbidities (15-17), sociodemographic information, comorbidities and current medications (19), or chest computed tomography (CT) scan with patient demographics (16). Some studies have also sought to predict prognosis by CT alone (19, 20). The performance of our tests compared favorably with that of two existing risk assessment models for which we were able to generate classifications for our development set, with our test achieving similar negative predictive values in the lowest risk groups but superior specificity. However, for other simple nomogram-based models, we were not able to compare performance due to lack of information for required attributes, as these were not obtainable from collected EHR data.

We utilized only attributes easily obtainable in an emergency care setting. This consideration is especially important given that many patients admitted with COVID-19 infection may not be in care and aware of comorbidities or may be critically ill upon presentation and unable to provide a detailed medical history. We selected simple clinical attributes (race, age, gender, weight) and augmented these with 22 readily available laboratory measures and vital signs, routinely collected at initial assessment, with results available within hours. This set of 26 attributes available in the EHR, proved sufficient to develop a suite of tests able to assess the prognosis for patients hospitalized with COVID-19 infection.

We selected disease severity measures of admission to the ICU, development of ARDS, and intubation as testable severe outcomes. This approach contrasts with some other studies (18) which have focused on predicting mortality, the rarity of which makes classifier training problematic, and which may be more strongly influenced by initial delays in seeking care, diagnosis, and more recent advances in treatment. Risk prediction of these shorter-term, more commonly occurring endpoints could be more relevant to guide discussions with patients and family members concerning aggressiveness of care and desires for intubation, channeling of scarce hospital resources to patients who would need them most, identification of those most likely to benefit from certain interventions or enrollment in clinical trials, and to provide peace of mind for patients at lower risk categories.

We carried out a blinded, independent validation three tests by generating test classifications blinded to all outcome data, and the results were analyzed according to a prespecified statistical analysis plan. This validation study was possible using data directly extracted from the EHR, without any further curation, demonstrating that the data required for risk assessment was easily and readily accessible. Test performance was similar between the development and validation cohorts, illustrating the ability of the ML platform to generalize to unseen datasets. The successful validation of the performance of our models observed in test development shows that our selection of minimal attributes was rich in predictive information. We explored incorporating additional attributes into our modelling (data not shown) but did not observe a significant improvement in test performance, even when including comorbidities and symptoms (5-7,11,17,18).

As with all modern ML-based modeling, it was not *a priori* obvious which attributes would most inform the results for an individual patient. Recent work using Shapley values has directly addressed model explainability at an individual patient level (27); in other words, Shapley values can determine the importance and use of model attributes for making a patient’s outcome prediction. Our ML architecture made the calculation of exact Shapley values for all classifiers for 50 patients from the validation cohort computationally feasible without the need for commonly made approximations (28-29, supplement).

The Shapley value analysis found that, when used in combination by the ML, simple clinical measures of risk and disease severity (age, weight, oxygen saturation) and general biomarkers of inflammation and acute phase reactants (CRP, d-dimer, LDH) were important for risk assessment. CRP, d-dimer, and LDH have been linked to COVID-19 severity in other studies, likely reflective of a pronounced inflammatory, systemic immune response with heightened thromboembolic risk (30-31). Increasing age and body mass index are well-established risk factors for influenza severity (32), and may, in part, exacerbate the inflammatory response (33-34). While race/ethnicity was not found to play a key role in explaining risk classification in the Shapley value analysis, a white race was observed to contribute towards a lower risk classification, while all other choices did not contribute substantially to risk category (see figure E22 in supplement). These observations were consistent with the associations we observed when we compared patient characteristics between our risk subgroups (see supplement). However, the Shapley values, which assess the relative importance of each attribute in the generation of a test result for an individual patient, also revealed that our tests combined information across the 26 attributes in non-trivial ways, which varied between patients. Classification was not driven by just a small subset of the attributes and the attributes that explained patient classification differed from patient to patient. For some patients, all attributes had similar importance, while for others, some attributes pointed towards higher risk, while others pointed towards lower risk.

One limitation of this study is the relatively small and geographically restricted validation cohort. While patients excluded from the development cohort due to missing data were generally similar to those included, in validation, patients with complete data (only 15% of the total available) exhibited higher rates of severe disease and generally worse prognostic factors (laboratory and vital signs) than those without. Further validation of the test in larger cohorts derived from other health systems and geographic areas is necessary.

In summary, we have developed and validated a suite of tests able to assess the risk of a poor outcome for patients hospitalized with COVID-19 based on information easily and routinely collected at time of hospital admission. Additional validation, preferably in a prospective setting, is required to further demonstrate the clinical utility of this risk assessment tool beyond clinical assessment alone. However, with readily-derived and quickly-available EHR data, a risk assessment at or near the time of admission can inform prognosis, guide discussions on the risks and benefits of treatments (including intubation), or identify low or high-risk patients for limited resources or enrollment in clinical trials. Furthermore, the methods here may be implemented in the care of future patients with novel viral infections.

## Supporting information

Results Supplement

TRIPOD Checklist

## Data Availability

Data and trained models may be made available upon request to the corresponding author

