## Supplementary material for "Predicting Prognosis in COVID-19 Patients using Machine Learning and Readily Available Clinical Data": Methods Supplement

### Supplement: Methods

#### Data Collection

The training data cohort contained patients aged 18 or older admitted to the University of Colorado Hospital patients with positive CoVID-19 nasopharyngeal polymerase chain reaction (PCR) between mid-March 2020 and mid-May 2020. Patient information was extracted from electronic medical records by medical students and stored in a REDCap project (E1). Patient data was restricted to first recorded admission, and to the first recorded observations for laboratory values and vital measures.

Electronic medical record data from the University of Colorado Hospital System was extracted and delivered by the university's data warehouse. Inclusion criteria for this datamart was hospitalization in a University of Colorado health system facility in conjunction with a positive CoVID-19 test or diagnosis. Testing data was restricted to patients aged 18 to 90 who had a first admission occurring after mid-May 2020 through mid-September 2020 to avoid overlap with training cohort. Laboratory data was restricted to the first observed within 7 days of admission; vital measures were restricted to first observed within 24 hrs of admission. Values below assay thresholds were converted to ½ lower limit of detection (84 [0.3%] values), values above threshold were converted to the median of observed values higher than the threshold (43 [0.2%] values). Outcome extraction methods for intubation, ARDS and ICU admission were validated using the chart extraction data as a gold standard for patients in the training set. Results are shown in Table E1.

**Table E1: Performance of outcome variable extraction from HDC data (reference: Epic Chart Manual Review)**

| | Training Subset (N= 291) | | | | Testing Subset (N= 122) | | | |
|---|---|---|---|---|---|---|---|---|
| **Measure** | **Sensitivity** | **Specificity** | **PPV** | **NPV** | **Sensitivity** | **Specificity** | **PPV** | **NPV** |
| ICU admission | 0.972 | 0.964 | 0.977 | 0.955 | 0.94 | 1.0 | 1.0 | 0.930 |
| ARDS | 0.939 | 0.929 | 0.792 | 0.981 | 0.971 | 0.921 | 0.825 | 0.988 |
| Intubation | 0.991 | 0.988 | 0.995 | 0.976 | 1.0 | 1.0 | 1.0 | 1.0 |

#### Classifier Development

All component classifiers developed for this study used Biodesix's Diagnostic Cortex® machine learning platform described in (E2). The nominal Diagnostic Cortex is an ensemble method for classification problems that incorporates elements from both traditional and modern machine learning such as information abstraction, boosting, bagging, and dropout regularization. Input attributes to the model are abstracted by first training a series of kNN classifiers that look only at small subsets of attributes to predict the classification label of interest. The predictions of these kNN classifiers are combined in a strongly dropped-out logistic regression (boosting) to get a final classification label for each sample. This is all done in a bagging scheme where the training data is sampled without replacement allowing for ensemble predictions and reliable performance estimates on the development set (out of bag estimates).

**Diagnostic Cortex Parameters:**

- 625 Train/Test realizations (bags) were used.
- The atomic classifiers were k-NN's with k=11 and were allowed to go up to 3 features deep, considering classifiers at all levels. No atomic classifier filtering was used.
- Standard logistic regression with dropout was used with 10 atomic classifiers left in at each of the 100,000 dropout iterations.

Some of the classifiers developed for this study used a novel extension of the Diagnostic Cortex platform where additional decision trees are used to incorporate additional clinical data. Instead of the nominal single split of the data into a single training and test set for each bag (training/test set realization), the data was split at two different points. First, the data is split to generate Training Set 1, for the nominal Master Classifier that combines the filtered atomic classifiers in a dropout logistic regression, and a reserve set. Then the reserve set is further sampled (30 times for this study) to train (Training Set 2) and evaluate (Test Set), with a set of random classification tree classifiers combining the output of the Master Classifier with clinical data for samples in Training Set 2. Samples in the evaluation Test Set are not used at all in training and can be used to generate out-of-bag classifications. This addition to the Diagnostic Cortex procedure is shown in Figure E1. For this study, the emergency department and basic bloodwork numeric features are used to train the nominal Diagnostic Cortex master classifiers, and the basic patient characteristics are combined with the master classifier output in the decision trees.

Referring to Figure E1, in each sampling of the reserved set, a classification tree was grown on Training Set 2 and the test set was evaluated with it. The data was split into 1/3 for Training Set 1, 2/3 for reserve, and then equal proportions of the subsequent splits of the reserve to Training Set 2 and the test set. In all samplings, an equal number of patients with the endpoint of interest were kept in each group.

The attributes passed to each tree were: the output of the Diagnostic Cortex Master Classifier (binarized with a cutoff at 0.5 and treated as categorical), age, gender, and race. Interaction terms were calculated between the binarized Diagnostic Cortex output and each of gender and race and passed to the trees as additional attributes. These interaction terms consist of categories representing each observed combination of the categories in the variables used to calculate them (eg gender = 'male' and Master Classifier output = 0)

The procedure for growing the classification trees for this study was as follows:

1. For non-categorical features, the values of each feature were sampled to get a list of feature values to consider when splitting each node in the tree. The values were taken to be the 10$^{th}$, 20$^{th}$, …, 90$^{th}$ percentile of observed feature values in the training set.
2. The training set was recursively split: The entire training set was assigned to the initial node. The procedure for each recursive node split was as follows:
    a. Sample the features in the training set: Take a random subset of *floor(sqrt(<number of features>))* features out of the list of the total set of features in the training data.
    b. If the depth of the tree exceeds 100: stop this recursion.
    c. For each possible value of the feature, for each feature in this feature subset, split the samples at this node by feature value into two candidate children nodes. If both groups have at least one sample present, consider this split valid. Calculate and store the weighted change in cross entropy from the parent node to the candidate children nodes. The class entropy for a set of samples with some classes is defined in terms of the class proportions in the set, $p_i$, as: $I_C = -\sum_{i \in classes} p_i * \log(p_i)$, with $0 * \log(0)$ taken to be the limit from the right which is $0$. The average entropy gain from the parent to the child nodes with the left node containing fraction $f_{left}$ of the samples and the right node containing fraction $f_{right}$ is: $L_C = I_C(parent) - (f_{left} * I_C(left) + f_{right} * I_C(right))$.
    d. If no possible splits are found, stop this recursion.
    e. Pick the feature and associated feature value that gives the largest change in cross entropy as defined in (c).
    f. Assign each group the label (event or no event) corresponding to the majority of its members.
    g. Split one child node.
    h. Split the other child node.

The trained classification trees now play the role of the logistic regression master classifiers in the original Diagnostic Cortex procedure for the purpose of obtaining the final label for a new sample. Out of bag estimates are obtained by looking at the prediction of each classification tree for which a sample was in the test set. For this study, a modified majority vote was used.

A set of binary classifiers were trained to directly predict 4 endpoints: admission to the ICU, development of any complication during hospitalization, development of acute respiratory distress syndrome during hospitalization, patient was intubated. All 4 cases used identical Diagnostic Cortex and Decision tree parameters as outlined above.

**Figure E1: The Diagnostic Cortex with Trees for Adding Additional Attributes**

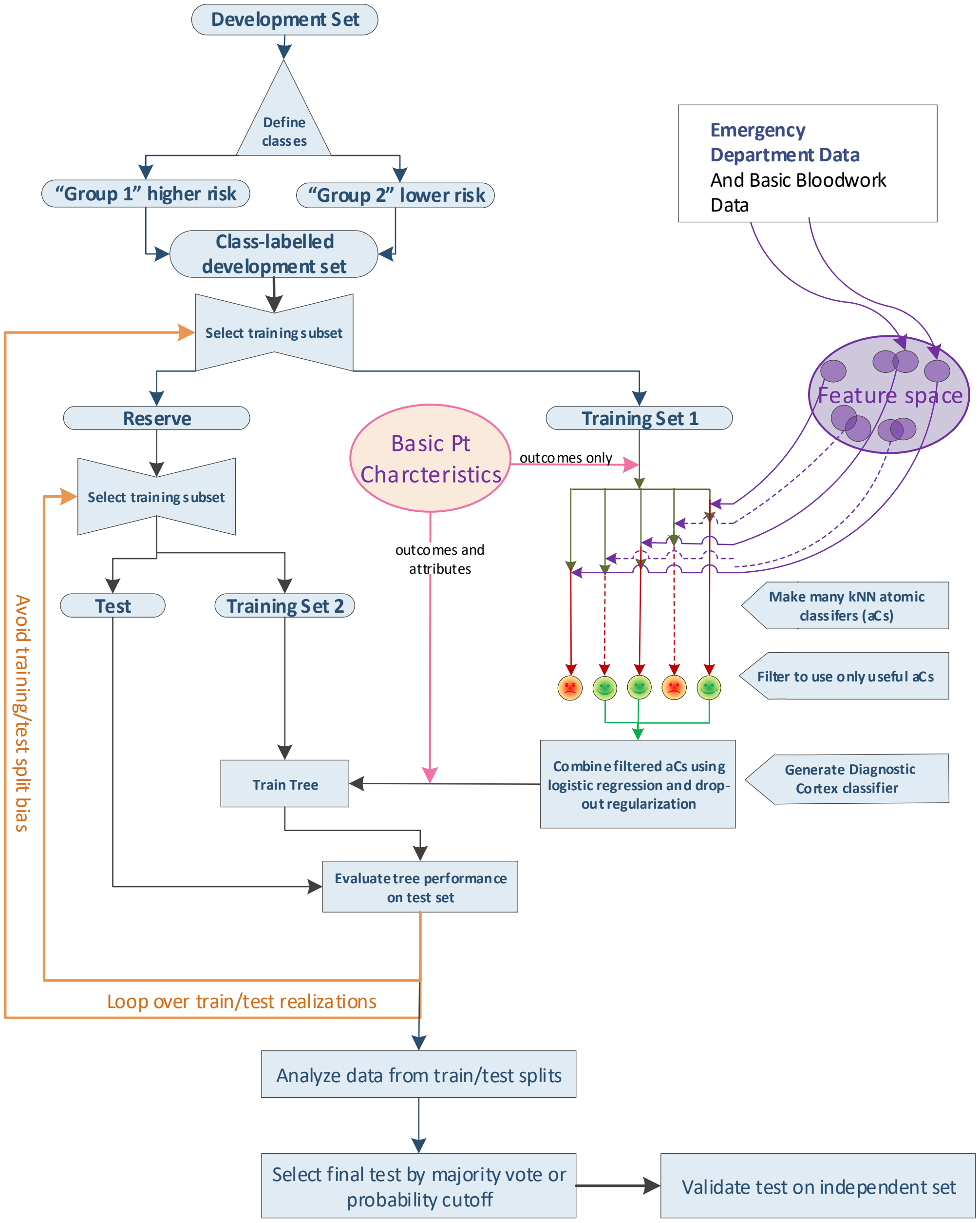


Additional (no-tree) Diagnostic Cortex models were trained to split the resulting groups of the binary classifiers in a hierarchical fashion. The resulting groups of the binary classifiers formed the development sets of the child classifiers. All models took a default cutoff value of 0.5 to assign patients either a ‘higher risk’ or ‘lower risk’ binary classification. All child models did not use trees and were trained only using the emergency department and bloodwork numbers. The Diagnostic Cortex operating parameters were the same as for the binary classifiers previously described. Two hierarchical configurations were used and are illustrated in figures E2 and E3.

**Figure E2: Hierarchical Configuration without an Intermediate Split:** A Diagnostic Cortex with in-bag decision tree model is used to split the entire development set into a higher and lower risk group for the endpoint. Diagnostic Cortex models without trees are used to split the two groups from the first split into four: lowest, low, high, highest.

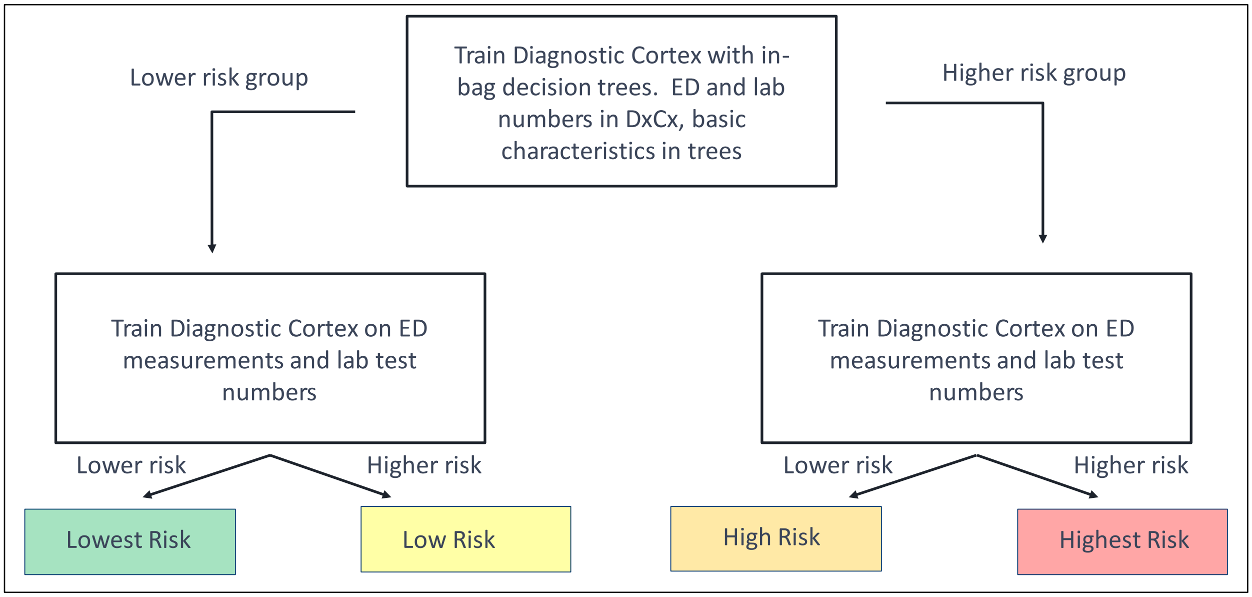


**Figure E3: Hierarchical Configuration with an Intermediate Split:** A Diagnostic Cortex with in-bag decision tree model is used to split the entire development set into a higher and lower risk group for the endpoint. Diagnostic Cortex models without trees are used to split the two groups from the first split into four. The lowest risk group is defined as a low risk classification by the first classifier and then a low risk classification by the second low risk classifier, and similar for the highest risk group. The other two groups are recombined and then another Diagnostic Cortex model is used to split this group into the final high and low risk groups.

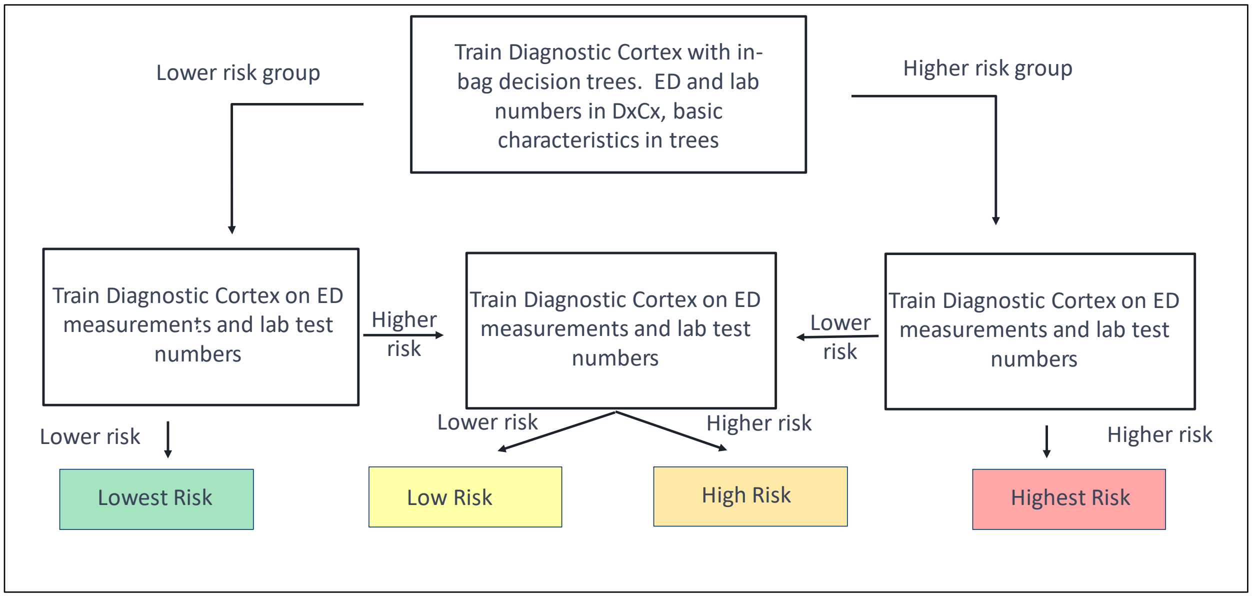

#### Shapley Values

The COVID risk assessment tests are good examples of how machine learning can be used to combine many patient attributes to produce a test able to predict a future clinical outcome with sufficient accuracy to provide clinical utility to physicians and patients. While the predictive accuracy of tests developed with modern machine learning (ML) can be high, the approach lacks transparency into the biological or clinical rationale underlying the test and does not provide a simple explanation of how the result for an individual patient is obtained from the input attributes.

Explainability of machine learning models and artificial intelligence has recently become a major focus of research. Concerns about biases in ML implementations, including those based on gender or race, and the recognition of the right of people to understand how their personal data is being used have focused attention on how to provide simple explanations and quantification of how attributes are used by complex ML algorithms. The EU General Data Protection Regulation of 2016 [E3] addresses this issue, providing individuals with the right to receive explanations of automated decisions carried out on the basis of their personal data.

Explainability research has moved away from trying to provide a global description of relative attribute importance accurate for all possible sets of input data, which is unlikely to be successful, given the complexity and nonlinearities inherent in most ML approaches, to a local approach of assessing relative attribute importance for the result for a specific set of input data. In the case of molecular tests, this latter approach corresponds to providing a description of the relative importance of the attributes used to the result generated for an individual patient.

One proposal has been to construct a simpler, more interpretable model of a complex ML algorithm which reproduces the results of the full ML algorithm in the vicinity of the particular result we seek to explain. The local interpretable model-agnostic explanation (LIME) approach [E4] seeks to generate a model of a ML algorithm which can be easily interpreted and which can reproduce the results of the ML algorithm in the locality of the result to be explained without requiring information about the ML algorithm itself. Only oracle access to the ML algorithm is required – i.e., only results of the ML algorithm for various inputs are necessary and no details of how they are arrived at from the input attributes. However, this approach relies on being able to train simpler, interpretable, locally accurate models in the vicinity of a result to be explained. In the case of binary or categorical tests, there are often large regions in feature space with consistent classifications (e.g. regions of feature space within which all feature values produce the same classification). Hence, locally only one test result can be generated. However, examples of both binary classes or multiple categorical classes are required to train the LIME model and these often do not exist in any sense locally to the sample for which we are seeking to explain the classification. Additionally, many ML models can demonstrate discontinuous behaviors, e.g., change in classification at a decision boundary for a k-nearest neighbor classifier, making it hard or impossible to define locality.

An alternative approach to explainability makes use of game theory concepts developed for determining the equitable distribution of winnings between players working in teams. Parallels between deciding on a fair distribution of winnings between team members in multiplayer games and assessing the relative importance of multiple input attributes to the result of a ML algorithm were observed several years ago. It was proposed that Shapley Values (SVs) [E5], which provide an equitable scheme for dividing game winnings within a team of multiple players, could provide the framework for assigning relative importance of multiple attributes to the result of a ML algorithm [E6,E7]. SVs assess the contribution of a player to the team's result, or the contributions of an attribute to the algorithm output, by examining the results/algorithm predictions for all possible coalitions of players within the team, or all possible subsets of attributes. Formally, let us assume that we have a predictor $f(\{S\})$ which depends on a set of attributes $\{S\}$. Further, we assume that we can define the prediction for any subset of attributes, $\{S\}$, contained within a set of all available attributes $\{M\}$.The SV for an attribute $j$ contained in $\{M\}$ is

$$\emptyset_j = \sum_{\{S\}\subseteq\{M\}\backslash\{j\}} \frac{|S|!\,(|M|-|S|-1)!}{|M|!}\,\left(f(\{S\}\cup\{j\}) - f(\{S\})\right)$$

SVs satisfy several axioms which guarantee that SV explanations have some properties that are logically consistent and aligned with human intuition. These properties include: efficiency (the entirety of the prediction result is distributed among the attributes), symmetry (attributes contributing in an equal manner are guaranteed to be assigned equal SVs), dummy player (attributes with definitively zero contribution to classification are assigned a SV of 0), linearity (the SVs for a prediction defined as the sum of two separate predictions is the sum of the SVs calculated individually for the two separate predictions) [E8,E9].

Unfortunately, the exact evaluation of SVs for ML algorithms using more than a handful of attributes is generally unfeasible. There are two main issues that arise:

1. The requirement to sum over all possible subsets of attributes

   This is a task of exponential complexity. For a set of attributes of size $|M|$, there are $2^{|M|}-1$ subsets that we have to sum over to find the SV for each member of $\{M\}$.

2. The need to be able to calculate the prediction of the complex ML model using only a subset of attributes.

   This problem is multi-faceted. First, one needs to be able to define what we mean by a prediction from the complex ML model with a smaller subset of features. Take a simple example of a k-nearest neighbor algorithm based on eight features. The direct interpretation of the ML model based on a subset of the eight features, would be a k-nearest neighbor algorithm trained on the same reference set as the original 8-feature model, using the same k, but using the smaller subset of features. As a k-nearest neighbor algorithm does not require a training phase, this could be implemented, provided that the classifier reference set were available. (Note that with only oracle access, this would not be possible, even for this simple example.) However, many complex ML algorithms require training to define model parameters and this would need to be repeated for each possible subset of attributes. This would quickly become compute-time prohibitive for complex models with even small numbers of attributes and even for simpler models that use larger numbers of attributes.

Several approaches have been proposed to alleviate these two issues. Kernel SHAP [E8] and extensions [E9], have been proposed to attempt to circumvent the need to retrain models on attribute subsets. However, these approaches break the correlation structures between attributes, lead to violation the exact SV axioms and can lead to results that can be counterintuitive and misleading [E9,E10]. Furthermore, they are not *approximations* to the exact SVs and there are no methods to understand how close the results are to the exact SVs. The first issue, summation of an exponential number of attribute subsets, has been addressed using sampling methods [E5,E8,E11]. These approaches are based on the observation that not all attribute subsets contribute equally to the Shapley sum and hence, sampling those subsets that contribute most at the expense of subsets contributing little, can lead to an approximation to the exact SV in less than exponential time. Monte Carlo-based sampling schemes should produce well-controlled approximations to exact SVs, although very little work has been done to characterize the convergence of these methods to exact SVs.

The ML approach used in development of the COVID risk stratification tests has particular properties that aid in evaluating SVs and hence providing explainability of individual test predictions.

The Diagnostic Cortex structure, composed of an average over many dropout iterations each of which is a coalition of a subset of the total number of available attributes, means that an exact prediction for the Diagnostic Cortex algorithm trained on the same reference set using a smaller number of attributes can be obtained *without the need for classifier retraining*. This will be explained in detail in a separate report. However, the principle is simple: one observes that a dropout iteration that is a coalition of a subset of features, $\{S\}$, created and trained for the model using the entire set of features $\{M\}$, is also an allowable dropout iteration for any subset of $\{M\}$ that includes $\{S\}$. This observation removes the need for classifier retraining (point 2 above) and so allows the calculation of approximations to SVs using Monte Carlo-based sampling to address point 1. Hence, we were able to generate SVs for the Diagnostic Cortex portions of the risk assessment classifiers using the sampling method of (E11) With 500,000 Monte Carlo sampling steps.

The remaining parts of the risk assessment classifiers were ensemble averages of trees constructed using only a very small number of attributes. The small number of attributes allowed an exact SV calculation over the exponential number of terms in the sum, with model retraining for each attribute subset.

SVs were evaluated for each of the classifiers used in each of the risk assessment tests for 50 patients from the validation cohort. Patients were selected so that there was somewhat equal representation across all possible test risk groups and endpoints. Race and gender were also considered in the selection, but representative populations across these attributes was secondary to risk group and endpoint.

### Statistical Analysis Plan for Validation of Suite of COVID Tests

10/15/2020

This document outlines the preplanned statistical analyses to be carried out as a validation of a suite of tests designed to identify risk of outcomes for hospitalized patients with COVID-19. Clinical data, blinded to all outcome, was provided by CU. Test classifications were generated and returned to CU on 10/15/2020. Outcome data will be provided only after confirmed receipt by CU of the blinded test classifications.

**Tests to be Validated**

Each test stratifies patients into groups of varying risk for a particular endpoint: admission to the ICU, development of any complication, development of ARDS, and need for intubation. The ICU, intubation, and any complication tests have four final risk groups: lowest risk, low risk, high risk, and highest risk. The ARDS classifier has only three final risk groups: lowest risk, high risk, and highest risk. Endpoint data for this validation analysis is only expected for ICU admission, intubation, and development of ARDS, but labels for risk of developing any complication will still be provided incase that endpoint data proves to be extractable at a later date.

**General Considerations**

*Study Population*

The study population includes all patients hospitalized with COVID-19 for whom test classifications could be generated from data extracted from UC Health Data Compass and received on 10/09/2020 and for whom at least one of the endpoints (i) admission to ICU (yes/no) and (ii) intubation (yes/no) is provided after unblinding.

*Statistical Considerations*

The statistical analyses contained in this SAP will be carried out using SAS (SAS9.4 or higher), GraphPad PRISM (PRISM 8 or higher), or Matlab2020. All analyses will be verified by an independent analyst. The results of all analyses will be contained in a final report containing this SAP as an appendix.

Additional ad hoc analyses may be performed and may be included in the final report, but will be cleared marked as outside the scope of this SAP.

**Analyses to be Performed**

*Primary Analysis*

1. The primary analysis to validate the performance of each test in the study population will be a test of trend of association of categorical test classification with endpoint. This will be carried

out for each predictive test separately with a prespecified significance level of $\alpha=0.05$ one-sided using the Cochran-Armitage test.

*Additional Analyses and Summary Statistics*

1. The clinical characteristics of the study population will be summarized, using frequency and percentages for categorical variables and mean, median, standard deviation and range for continuous variables.
2. The clinical characteristics of the study population will be summarized by test classification for each test, using frequency and percentages for categorical variables and mean, median, standard deviation and range for continuous variables. Association of test classification with clinical characteristics will be assessed using Mann-Whitney tests, Fisher's exact tests (when 3 or fewer categories) or chi-squared tests (when 4 or more categories).
3. The association of test classification of highest risk vs other and lowest risk vs other with outcome endpoint will be assessed using Fisher's exact test.
4. The percentage of patients positive or negative for each endpoint will be calculated for each test classification along with associated 95% confidence intervals. The precision and recall will be calculated for each test classification group.
5. The positive predictive value will be calculated for the highest risk test classification vs the other test classification groups.
6. The negative predictive value will be calculated for the lowest risk test classification vs the other test classification groups.
7. The areas under the receiver operating characteristic (ROC) curve will be calculated for the continuous variables generated during test classification generation.
