## Supplementary material for "Predicting Prognosis in COVID-19 Patients using Machine Learning and Readily Available Clinical Data": Results Supplement

### Supplement: Results

#### Patient Cohorts

For patients to be included in either of the development or validation cohorts, all necessary attributes had to be complete. This requirement led to 229 patients being included in the development cohort and 222 excluded from the available 451 patients. In the validation cohort, 330 patients were included and 1,933 were excluded from the available 2,263. Tables E2 and E3 compare the categorical and continuous attributes (including endpoints) used in training the classifiers between the included patients and the excluded patients in development. Tables E4 and E5 provide the same comparisons in validation. For the categorical attributes and endpoints, the number of patients per category and the percent contribution of each category are given for the included and excluded patients. P-values for Fisher's exact test of proportions between the included and excluded sets of patients are also given. For the continuous attributes, the median and inter-quartile range are given for the included and excluded patients. P-values for Mann-Whitney tests comparing the attribute distributions between the included and excluded patients are also given. In addition to the p-values, a measure of effect size is given for both the categorical and continuous attributes. Cohen's w (categorical) and d (continuous) have the rule-of-thumb interpretations of 0.2 or less being a small effect size and greater than 0.8 being a large effect size for absolute value Cohen's d and 0.1 or less being a small effect size and greater than 0.5 being a large effect size for absolute value Cohen's w [E12]. Effect sized categorized as large by these rules are marked in bold. In calculating the effect size measures, any attribute that was found to be non-normally distributed by a Kolmogorov-Smirnov test with a confidence level of 0.01 was log transformed before calculating the effect size.

**Table E2: Comparison of categorical attributes and endpoints in development**

| Attribute | Class | Included | Excluded | Cohen w | Fisher p |
|---|---|---|---|---|---|
| Race | | | | 0.2115 | 0.278 |
| | NA | 0 (0%) | 0 (0%) | | |
| | White | 41 (17.9%) | 56 (25.23%) | | |
| | Black/African American | 52 (22.71%) | 51 (22.97%) | | |
| | Hispanic/Latino | 94 (41.05%) | 80 (36.04%) | | |
| | Other | 31 (13.54%) | 29 (13.06%) | | |
| | Unknown | 11 (4.803%) | 6 (2.703%) | | |
| Gender | | | | 0.00715 | 0.962 |
| | NA | 0 (0%) | 0 (0%) | | |
| | Male | 124 (54.15%) | 121 (54.5%) | | |
| | Female | 105 (45.85%) | 101 (45.5%) | | |
| eGFR | | | | **0.6226** | 0.0889 |
| | NA | 0 (0%) | 94 (42.34%) | | |
| | x>=60 | 180 (78.6%) | 96 (43.24%) | | |

| | | | | | |
|---|---|---|---|---|---|
| | 30>=x>60 | 34 (14.85%) | 15 (6.757%) | | |
| | x<30 | 15 (6.55%) | 17 (7.658%) | | |
| ICU Admission | | | | 0.1587 | 0.0982 |
| | NA | 0 (0%) | 0 (0%) | | |
| | Yes | 77 (33.62%) | 92 (41.44%) | | |
| | No | 152 (66.38%) | 130 (58.56%) | | |
| Intubated | | | | 0.225 | 0.0161 |
| | NA | 0 (0%) | 0 (0%) | | |
| | No | 176 (76.86%) | 147 (66.22%) | | |
| | Yes | 53 (23.14%) | 75 (33.78%) | | |
| ARDS Diagnosis | | | | 0.1379 | 0.144 |
| | NA | 0 (0%) | 0 (0%) | | |
| | 0 | 184 (80.35%) | 165 (74.32%) | | |
| | 1 | 45 (19.65%) | 57 (25.68%) | | |

**Table E3: Comparison of continuous attributes in development**

| Attribute | Group | number NA | Median (IQR) | Mann-Whitney p | Cohen d |
|---|---|---|---|---|---|
| Age, years | | | | 0.3042 | 0.09093 |
| | Excluded | 0 | 54.5 (41.0 – 67.0) | | |
| | Included | 0 | 57 (43.0 – 68.0) | | |
| Temperature, °C | | | | 0.4602 | -0.006523 |
| | Excluded | 1 | 37.0 (36.5 - 37.7) | | |
| | Included | 0 | 36.9 (36.5 - 37.5) | | |
| Heart Rate, beats/minute | | | | 0.1448 | 0.1488 |
| | Excluded | 0 | 96.5 (84.0 – 109.0) | | |
| | Included | 0 | 98.0 (84.0 - 113.3) | | |
| Systolic BP, mm Hg | | | | 0.01135 | 0.2381 |
| | Excluded | 0 | 128 (115 - 141) | | |
| | Included | 0 | 132 (118 - 146) | | |
| Diastolic BP, mm Hg | | | | 0.3625 | 0.06175 |
| | Excluded | 0 | 72 (65 - 81) | | |
| | Included | 0 | 74 (65 - 83) | | |
| Respiratory Rate, breaths/minute | | | | 0.2947 | 0.09679 |
| | Excluded | 2 | 20 (18 - 24) | | |

| | | | | | |
|---|---|---|---|---|---|
| | Included | 0 | 20 (18 - 24) | | |
| Oxygen Saturation, % | | | | 0.02695 | -0.14066 |
| | Excluded | 1 | 92 (90 - 95) | | |
| | Included | 0 | 92 (87 - 94) | | |
| Weight, kg | | | | 0.2603 | 0.08343 |
| | Excluded | 2 | 81.6 (69.5 – 94.5) | | |
| | Included | 0 | 81.6 (72.0 - 99.8) | | |
| QTc | | | | 0.3946 | -0.09557 |
| | Excluded | 19 | 442 (423 - 462) | | |
| | Included | 0 | 440 (422 - 459) | | |
| Sodium, mmol/L | | | | 0.2226 | 0.09264 |
| | Excluded | 87 | 135 (133 - 138) | | |
| | Included | 0 | 136 (134 - 138) | | |
| Potassium, mmol/L | | | | 0.2769 | -0.21914 |
| | Excluded | 87 | 3.8 (3.4 - 4.2) | | |
| | Included | 0 | 3.8 (3.5 – 4.0) | | |
| Carbon Dioxide, mmol/L | | | | 0.1348 | 0.282 |
| | Excluded | 87 | 22 (20 - 24) | | |
| | Included | 0 | 23 (21 - 25) | | |
| BUN, mg/dL | | | | 0.127 | -0.17709 |
| | Excluded | 87 | 15 (11 - 26) | | |
| | Included | 0 | 13 (10 - 20) | | |
| Creatinine, mg/dL | | | | 0.153 | -0.24844 |
| | Excluded | 87 | 0.96 (0.74 - 1.28) | | |
| | Included | 0 | 0.94 (0.70 - 1.20) | | |
| Anion Gap, mmol/L | | | | 0.2101 | -0.10248 |
| | Excluded | 92 | 12 (10 - 14) | | |
| | Included | 0 | 12 (10 - 13) | | |
| WBC Count, 109 cells/L | | | | 0.2531 | -0.09721 |
| | Excluded | 87 | 7.1 (5.2 - 10.0) | | |
| | Included | 0 | 6.8 (5.3 - 8.9) | | |
| Hemoglobin, g/dL | | | | 0.1543 | 0.1768 |
| | Excluded | 89 | 14.0 (12.3 - 15.9) | | |
| | Included | 0 | 14.6 (13.3 - 15.7) | | |
| Hematocrit, % | | | | 0.1145 | 0.165 |
| | Excluded | 87 | 42.0 (37.8 - 47.3) | | |
| | Included | 0 | 43.7 (39.8 - 46.8) | | |

| | | | | | |
|---|---|---|---|---|---|
| Platelet Count, 109 cells/L | | | | 0.1387 | 0.09887 |
| | Excluded | 88 | 199 (148 - 240) | | |
| | Included | 0 | 212 (162 - 263) | | |
| LDH, U/L | | | | 0.2516 | -0.14958 |
| | Excluded | 156 | 338 (263 - 476) | | |
| | Included | 0 | 317 (258 - 422) | | |
| D-Dimer ng/mL | | | | 0.02823 | -0.35868 |
| | Excluded | 146 | 945 (660 - 2500) | | |
| | Included | 0 | 860 (530 - 1453) | | |
| CRP, mg/L | | | | 0.8575 | 0.01235 |
| | Excluded | 115 | 89.4 (36.9 - 161) | | |
| | Included | 0 | 83.3 (40.9 - 146) | | |
| Ferritin, ng/mL | | | | 0.8255 | 0.02979 |
| | Excluded | 164 | 341 (186 - 759) | | |
| | Included | 0 | 364 (170 - 726) | | |

**Table E4: Comparison of categorical attributes and endpoints in validation**

| Attribute | Class | Included | Excluded | Cohen w | Fisher p |
|---|---|---|---|---|---|
| Race | | | | **0.78** | <0.0001 |
| | NA | 0 (0%) | 0 (0%) | | |
| | White | 113 (34.24%) | 1290 (66.74%) | | |
| | Black/African American | 23 (6.97%) | 112 (5.794%) | | |
| | Hispanic/Latino | 164 (49.7%) | 386 (19.97%) | | |
| | Other | 25 (7.576%) | 127 (6.57%) | | |
| | Unknown | 5 (1.515%) | 18 (0.9312%) | | |
| Gender | | | | 0.3779 | <0.0001 |
| | NA | 0 (0%) | 0 (0%) | | |
| | Male | 172 (52.12%) | 661 (34.2%) | | |
| | Female | 158 (47.88%) | 1272 (65.8%) | | |
| eGFR | | | | 0.3128 | 0.152 |
| | NA | 0 (0%) | 473 (24.47%) | | |
| | x>=60 | 244 (73.94%) | 1150 (59.49%) | | |
| | 30>=x>60 | 61 (18.48%) | 222 (11.48%) | | |
| | x<30 | 25 (7.576%) | 88 (4.553%) | | |
| ICU Admission | | | | 0.3164 | <0.0001 |
| | NA | 0 (0%) | 0 (0%) | | |
| | Yes | 85 (25.76%) | 282 (14.59%) | | |

| | | | | | |
|---|---|---|---|---|---|
| | No | 245 (74.24%) | 1651 (85.41%) | | |
| Intubated | | | | 0.3037 | <0.0001 |
| | NA | 0 (0%) | 0 (0%) | | |
| | No | 288 (87.27%) | 1823 (94.31%) | | |
| | Yes | 42 (12.73%) | 110 (5.691%) | | |
| ARDS Diagnosis | | | | **0.6468** | <0.0001 |
| | NA | 0 (0%) | 0 (0%) | | |
| | 0 | 295 (89.39%) | 1897 (98.14%) | | |
| | 1 | 35 (10.61%) | 36 (1.862%) | | |

**Table E5: Comparison of continuous attributes in validation**

| Attribute | Group | number NA | Median (IQR) | Mann-Whitney p | Cohen d |
|---|---|---|---|---|---|
| Age, years | | | | <0.0001 | 0.3621 |
| | Excluded | 0 | 49 (33 - 66) | | |
| | Included | 0 | 57 (44 - 70) | | |
| Temperature, °C | | | | <0.0001 | 0.5279 |
| | Excluded | 110 | 36.7 (36.4 – 37.0) | | |
| | Included | 0 | 37.0 (36.6 - 37.6) | | |
| Heart Rate, beats/minute | | | | <0.0001 | 0.5112 |
| | Excluded | 109 | 86 (75 - 100) | | |
| | Included | 0 | 98 (85 - 111) | | |
| Systolic BP, mm Hg | | | | 0.123 | -0.09211 |
| | Excluded | 112 | 130 (118 - 142) | | |
| | Included | 0 | 127 (116 - 142) | | |
| Diastolic BP, mm Hg | | | | <0.0001 | -0.2581 |
| | Excluded | 112 | 78 (69 - 87) | | |
| | Included | 0 | 74 (65 - 84) | | |
| Respiratory Rate, breaths/minute | | | | <0.0001 | 0.5876 |
| | Excluded | 109 | 18 (16 - 20) | | |
| | Included | 0 | 20 (18 - 24) | | |
| Oxygen Saturation, % | | | | <0.0001 | -0.44266 |
| | Excluded | 109 | 96 (93 - 97) | | |
| | Included | 0 | 92 (88 - 95) | | |
| Weight, kg | | | | 0.001491 | 0.2186 |
| | Excluded | 218 | 81.7 (69.5 - 97.1) | | |
| | Included | 0 | 85.4 (72.6 - 104.4) | | |

| | | | | | |
|---|---|---|---|---|---|
| QTc | | | | 0.0001928 | -0.22254 |
| | Excluded | 1183 | 451 (431 - 476) | | |
| | Included | 0 | 442 (424 - 467) | | |
| Sodium, mmol/L | | | | 0.002981 | -0.16961 |
| | Excluded | 479 | 136 (134 - 139) | | |
| | Included | 0 | 136 (133 - 139) | | |
| Potassium, mmol/L | | | | 0.2369 | -0.03967 |
| | Excluded | 474 | 3.9 (3.6 - 4.2) | | |
| | Included | 0 | 3.9 (3.6 - 4.2) | | |
| Carbon Dioxide, mmol/L | | | | 0.8544 | -0.031904 |
| | Excluded | 479 | 23 (20 - 25) | | |
| | Included | 0 | 23 (21 - 25) | | |
| BUN, mg/dL | | | | 0.2261 | 0.08509 |
| | Excluded | 479 | 14 (10 - 20) | | |
| | Included | 0 | 15 (10 - 22) | | |
| Creatinine, mg/dL | | | | 0.00568 | 0.1259 |
| | Excluded | 473 | 0.82 (0.67 - 1.07) | | |
| | Included | 0 | 0.87 (0.72 - 1.16) | | |
| Anion Gap, mmol/L | | | | <0.0001 | 0.4234 |
| | Excluded | 489 | 10 (8 - 11) | | |
| | Included | 0 | 11 (9 - 13) | | |
| WBC Count, 109 cells/L | | | | <0.0001 | -0.53024 |
| | Excluded | 97 | 9.5 (7.0 - 12.4) | | |
| | Included | 0 | 6.95 (5.5 - 9.1) | | |
| Hemoglobin, g/dL | | | | <0.0001 | 0.4518 |
| | Excluded | 83 | 13.1 (11.6 - 14.3) | | |
| | Included | 0 | 14.1 (12.8 - 15.3) | | |
| Hematocrit, % | | | | <0.0001 | 0.489 |
| | Excluded | 70 | 39.0 (35.4 - 42.5) | | |
| | Included | 0 | 42.1 (38.3 - 45.5) | | |
| Platelet Count, 109 cells/L | | | | <0.0001 | -0.25436 |
| | Excluded | 95 | 223 (179 - 273) | | |
| | Included | 0 | 198 (157 - 255) | | |
| LDH, U/L | | | | 0.02186 | -0.16421 |
| | Excluded | 1640 | 529 (346 - 860) | | |
| | Included | 0 | 446 (285 - 753) | | |
| D-Dimer ng/mL | | | | 0.01495 | -0.18322 |

| | | | | | |
|---|---|---|---|---|---|
| | Excluded | 1569 | 905 (485 - 1937) | | |
| | Included | 0 | 737 (405 - 1450) | | |
| CRP, mg/L | | | | 0.488 | 0.07159 |
| | Excluded | 1497 | 71.4 (27.7 - 173.9) | | |
| | Included | 0 | 79.4 (36.6 - 159.9) | | |
| Ferritin, ng/mL | | | | 0.9159 | 0.01625 |
| | Excluded | 1729 | 318 (135 - 679) | | |
| | Included | 0 | 308 (126 - 602) | | |

#### Classifier Development

Note that all results for the development cohort were obtained using out-of-bag estimates [E13].

#### Prediction of ICU admission

Figures E4 through E7 compare the receiver-operating characteristic (ROC) curves in development and validation for the individual component classifiers in the ICU test with the cutoff value of 0.5 marked for each. Areas under the curve (AUCs) are also given in the legends. Tables E2 and E3 respectively summarize the categorical and continuous attributes used in prediction for each of the final risk groups of the ICU test in development. Tables E4 and E5 give the same in the validation set. Mann-Whitney p-values are given comparing the continuous attribute ranks between the highest and lowest risk groups compared to all other risk groups. All statistics in these tables were calculated using Matlab R2020a. In the validation cohort, for race, a chi-squared test of independence was calculated with race grouped as: white, black, Hispanic, and other. The p-value was 0.040. For EGFR and gender, a Fisher exact test was used, yielding p-values of 0.118 and 0.0002 respectively. These statistics were calculated using SAS Enterprise Guide 8.2 (SAS 9.4).

**Figure E4: ROC Curve Comparison for the First Split Classifier for ICU Admission**

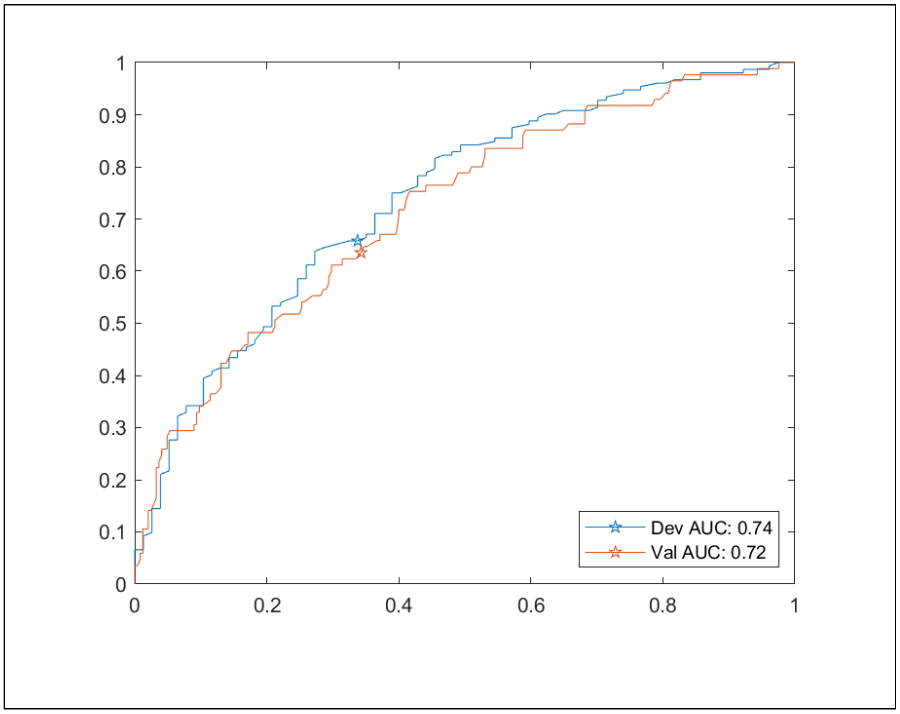

**Figure E5: ROC Curve Comparison for the Low Risk Child Classifier for ICU Admission**

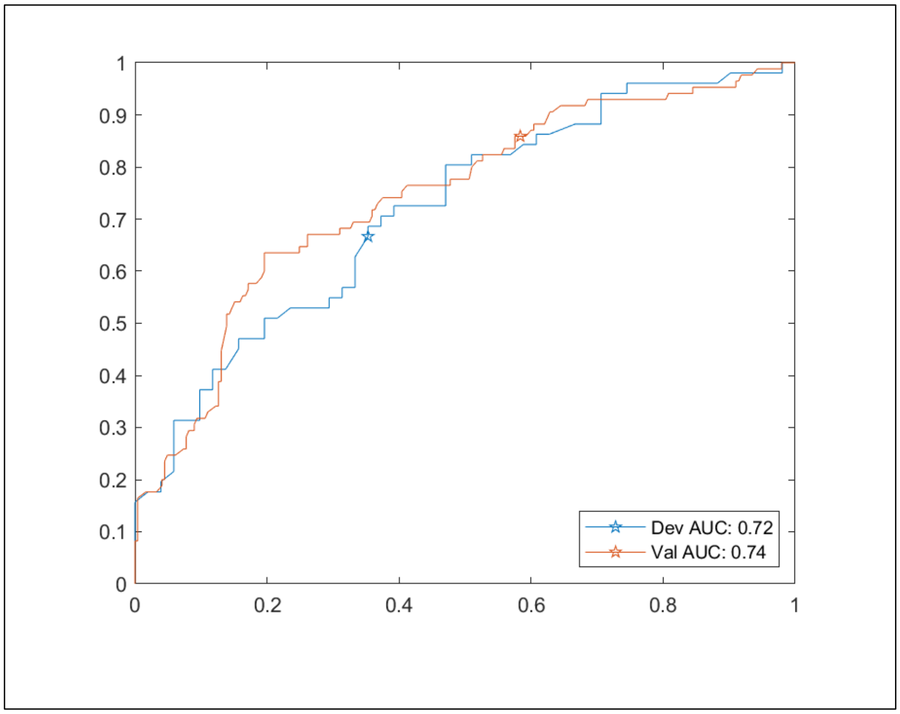


**Figure E6: ROC Curve Comparison for the High Risk Child Classifier for ICU Admission**

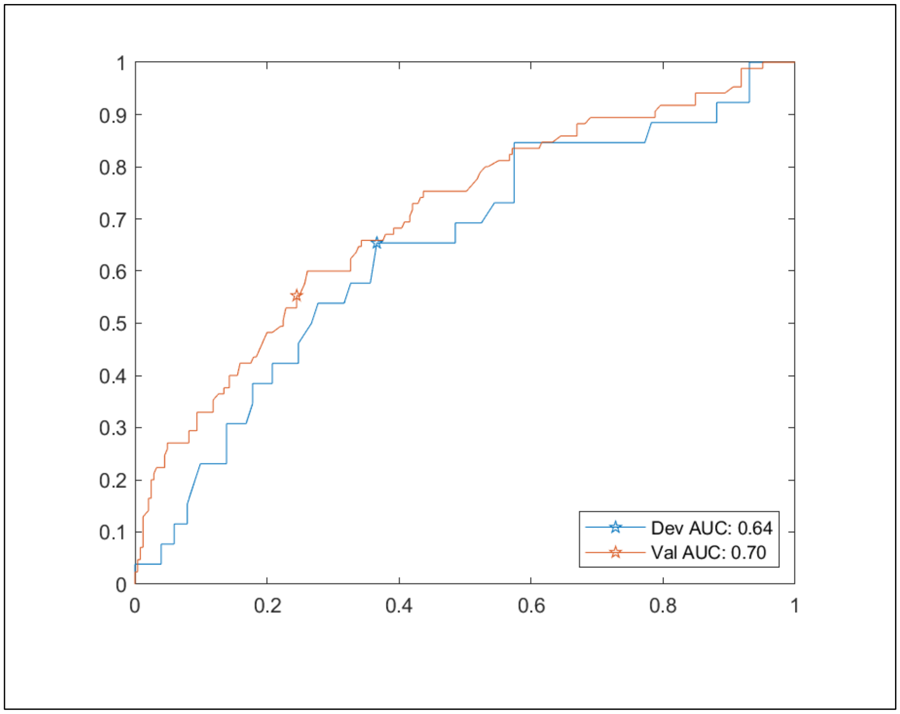

**Figure E7: ROC Curve Comparison for the Intermediate Risk Grandchild Classifier for ICU Admission**

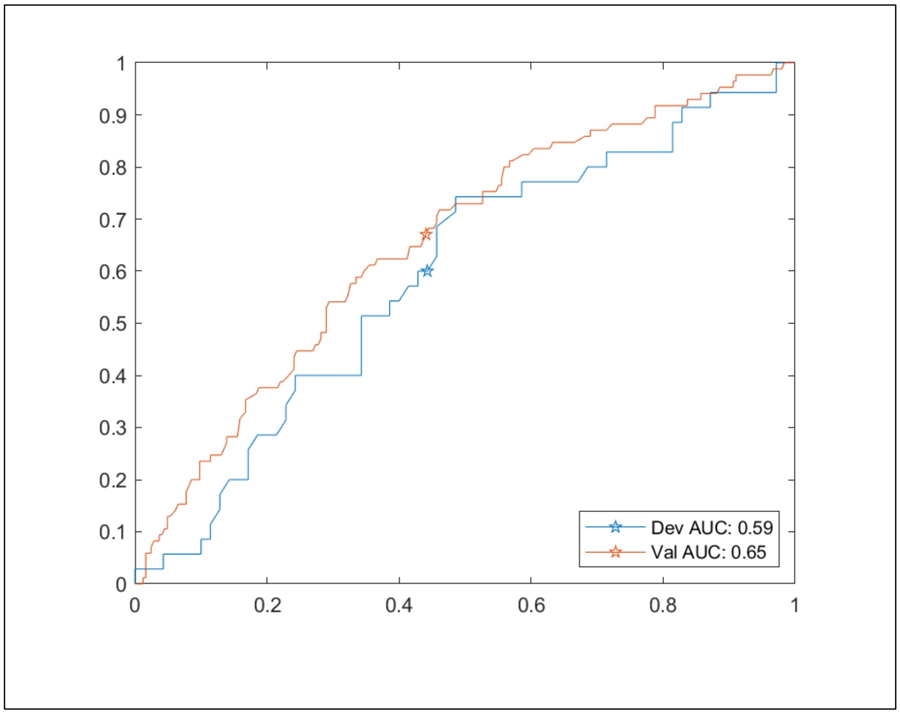


**Table E6: Summary of Categorical Attributes by ICU Admission Risk Group in Development**

| Attribute | Class | lowest | low | high | highest |
|---|---|---|---|---|---|
| | | n (proportion of group) | | | |
| Race | | N=73 | N=54 | N=51 | N=51 |
| | White | 21 (0.288) | 12 (0.222) | 6 (0.118) | 2 (0.039) |
| | Black/African American | 14 (0.192) | 13 (0.241) | 11 (0.216) | 14 (0.275) |
| | Hispanic/Latino | 31 (0.425) | 19 (0.352) | 23 (0.451) | 21 (0.412) |
| | Other | 6 (0.082) | 7 (0.130) | 8 (0.157) | 10 (0.196) |
| | Unknown | 1 (0.014) | 3 (0.056) | 3 (0.059) | 4 (0.078) |
| Gender | | N=73 | N=54 | N=51 | N=51 |
| | Male | 27 (0.370) | 34 (0.630) | 25 (0.490) | 38 (0.745) |
| | Female | 46 (0.630) | 20 (0.370) | 26 (0.510) | 13 (0.255) |
| eGFR | | N=73 | N=54 | N=51 | N=51 |
| | x>=60 | 67 (0.918) | 40 (0.741) | 44 (0.863) | 29 (0.569) |
| | 30>=x>60 | 5 (0.068) | 7 (0.130) | 4 (0.078) | 18 (0.353) |
| | x<30 | 1 (0.014) | 7 (0.130) | 3 (0.059) | 4 (0.078) |

**Table E7: Summary of Continuous Attributes by ICU Admission Risk Group in Development**

| Attribute | Label | Median | 25th Percentile | 75th Percentile | Mann-Whitney p |
|---|---|---|---|---|---|
| Age, years | | | | | |
| | lowest | 54 | 36 | 68 | 0.245 |
| | low | 60 | 49 | 69 | |
| | high | 57 | 40 | 65 | |
| | highest | 55 | 48 | 68 | 0.791 |
| Temperature, °C | | | | | |
| | lowest | 37 | 36 | 38 | 0.915 |
| | low | 37 | 37 | 37 | |
| | high | 37 | 36 | 37 | |
| | highest | 37 | 37 | 38 | 0.241 |
| Heart Rate, beats/minute | | | | | |
| | lowest | 110 | 86 | 110 | 0.339 |
| | low | 92 | 84 | 110 | |
| | high | 93 | 82 | 110 | |
| | highest | 110 | 89 | 120 | 0.003 |
| Systolic BP, mm Hg | | | | | |
| | lowest | 140 | 120 | 160 | 0.02 |
| | low | 130 | 120 | 150 | |
| | high | 120 | 110 | 130 | |
| | highest | 140 | 130 | 150 | 0.212 |
| Diastolic BP, mm Hg | | | | | |
| | lowest | 77 | 70 | 87 | 0.005 |
| | low | 73 | 64 | 79 | |
| | high | 66 | 60 | 80 | |
| | highest | 76 | 68 | 85 | 0.192 |
| Respiratory Rate, breaths/minute | | | | | |
| | lowest | 20 | 18 | 23 | 0.082 |
| | low | 20 | 18 | 24 | |
| | high | 18 | 18 | 22 | |
| | highest | 24 | 20 | 29 | < 0.001 |
| Oxygen Saturation, % | | | | | |
| | lowest | 93 | 91 | 95 | < 0.001 |
| | low | 92 | 88 | 94 | |
| | high | 91 | 86 | 94 | |
| | highest | 85 | 78 | 91 | < 0.001 |
| Weight, kg | | | | | |
| | lowest | 77 | 70 | 82 | < 0.001 |
| | low | 85 | 71 | 110 | |
| | high | 85 | 73 | 100 | |

| | | | | | |
|---|---|---|---|---|---|
| | highest | 94 | 76 | 110 | 0.008 |
| QTc | | | | | |
| | lowest | 450 | 420 | 460 | 0.366 |
| | low | 440 | 430 | 460 | |
| | high | 420 | 420 | 450 | |
| | highest | 440 | 430 | 470 | 0.279 |
| Sodium, mmol/L | | | | | |
| | lowest | 140 | 130 | 140 | 0.233 |
| | low | 140 | 130 | 140 | |
| | high | 140 | 130 | 140 | |
| | highest | 140 | 130 | 140 | 0.795 |
| Potassium, mmol/L | | | | | |
| | lowest | 3.7 | 3.4 | 3.9 | 0.038 |
| | low | 3.7 | 3.5 | 3.9 | |
| | high | 3.8 | 3.4 | 4 | |
| | highest | 3.9 | 3.6 | 4.2 | 0.014 |
| Carbon Dioxide, mmol/L | | | | | |
| | lowest | 23 | 21 | 25 | 0.082 |
| | low | 23 | 21 | 24 | |
| | high | 23 | 21 | 25 | |
| | highest | 22 | 20 | 24 | 0.023 |
| BUN, mg/dL | | | | | |
| | lowest | 12 | 8 | 15 | < 0.001 |
| | low | 13 | 12 | 25 | |
| | high | 13 | 10 | 17 | |
| | highest | 20 | 14 | 31 | < 0.001 |
| Creatinine, mg/dL | | | | | |
| | lowest | 0.69 | 0.6 | 0.9 | < 0.001 |
| | low | 1 | 0.79 | 1.3 | |
| | high | 0.95 | 0.72 | 1.1 | |
| | highest | 1.1 | 0.94 | 1.7 | < 0.001 |
| Anion Gap, mmol/L | | | | | |
| | lowest | 11 | 9.8 | 13 | 0.002 |
| | low | 12 | 10 | 13 | |
| | high | 12 | 10 | 13 | |
| | highest | 13 | 12 | 15 | < 0.001 |
| WBC Count, 109 cells/L | | | | | |
| | lowest | 5.8 | 4.3 | 7.1 | < 0.001 |
| | low | 6.8 | 5.6 | 8.5 | |
| | high | 7.1 | 5.3 | 8.5 | |
| | highest | 8.9 | 6.7 | 11 | < 0.001 |
| Hemoglobin, g/dL | | | | | |
| | lowest | 15 | 13 | 15 | 0.62 |

| | | | | | |
|---|---|---|---|---|---|
| | low | 15 | 13 | 16 | |
| | high | 14 | 13 | 15 | |
| | highest | 15 | 14 | 16 | 0.007 |
| Hematocrit, % | | | | | |
| | lowest | 44 | 39 | 46 | 0.666 |
| | low | 43 | 39 | 46 | |
| | high | 42 | 39 | 45 | |
| | highest | 46 | 41 | 49 | 0.003 |
| Platelet Count, 109 cells/L | | | | | |
| | lowest | 220 | 170 | 270 | 0.16 |
| | low | 210 | 170 | 240 | |
| | high | 210 | 150 | 270 | |
| | highest | 200 | 150 | 300 | 0.849 |
| LDH, U/L | | | | | |
| | lowest | 250 | 220 | 300 | < 0.001 |
| | low | 320 | 280 | 360 | |
| | high | 360 | 290 | 460 | |
| | highest | 440 | 360 | 580 | < 0.001 |
| D-Dimer ng/mL | | | | | |
| | lowest | 560 | 410 | 1000 | < 0.001 |
| | low | 840 | 410 | 1500 | |
| | high | 900 | 700 | 1500 | |
| | highest | 1200 | 800 | 2400 | < 0.001 |
| CRP, mg/L | | | | | |
| | lowest | 48 | 16 | 71 | < 0.001 |
| | low | 66 | 30 | 130 | |
| | high | 100 | 78 | 150 | |
| | highest | 150 | 100 | 250 | < 0.001 |
| Ferritin, ng/mL | | | | | |
| | lowest | 230 | 81 | 340 | < 0.001 |
| | low | 440 | 290 | 720 | |
| | high | 480 | 190 | 1000 | |
| | highest | 590 | 310 | 1200 | < 0.001 |

**Table E8: Summary of Categorical Attributes by ICU Admission Risk Group in Validation**

| **Attribute** | **Class** | **lowest** | **low** | **high** | **highest** |
|---|---|---|---|---|---|
| | | **n (proportion of group)** | | | |
| Race | | | | | |
| | White | 43 (0.457) | 21 (0.284) | 34 (0.386) | 15 (0.203) |
| | Black/African American | 7 (0.074) | 6 (0.081) | 5 (0.057) | 5 (0.068) |
| | Hispanic/Latino | 36 (0.383) | 37 (0.500) | 43 (0.489) | 48 (0.649) |

| | | | | | |
|---|---|---|---|---|---|
| | Other | 8 (0.085) | 9 (0.122) | 5 (0.057) | 3 (0.041) |
| | Unknown | 0 (0.000) | 1 (0.014) | 1 (0.011) | 3 (0.041) |
| Gender | | | | | |
| | Male | 41 (0.436) | 44 (0.595) | 35 (0.398) | 52 (0.703) |
| | Female | 53 (0.564) | 30 (0.405) | 53 (0.602) | 22 (0.297) |
| eGFR | | | | | |
| | x>=60 | 74 (0.787) | 54 (0.730) | 64 (0.727) | 52 (0.703) |
| | 30>=x>60 | 18 (0.191) | 14 (0.189) | 18 (0.205) | 11 (0.149) |
| | x<30 | 2 (0.021) | 6 (0.081) | 6 (0.068) | 11 (0.149) |

**Table E9: Summary of Continuous Attributes by ICU Admission Risk Group in Validation**

| Attribute | Label | Median | 25th Percentile | 75th Percentile | Mann-Whitney p |
|---|---|---|---|---|---|
| Age, years | | | | | |
| | lowest | 57 | 43 | 71 | 0.757 |
| | low | 59 | 42 | 71 | |
| | high | 59 | 49 | 72 | |
| | highest | 55 | 43 | 66 | 0.242 |
| Temperature, °C | | | | | |
| | lowest | 37 | 37 | 37 | 0.142 |
| | low | 37 | 37 | 37 | |
| | high | 37 | 37 | 38 | |
| | highest | 37 | 37 | 38 | 0.439 |
| Heart Rate, beats/minute | | | | | |
| | lowest | 94 | 82 | 110 | 0.049 |
| | low | 97 | 84 | 110 | |
| | high | 94 | 82 | 110 | |
| | highest | 110 | 96 | 120 | < 0.001 |
| Systolic BP, mm Hg | | | | | |
| | lowest | 130 | 120 | 150 | 0.115 |
| | low | 130 | 120 | 140 | |
| | high | 120 | 110 | 130 | |
| | highest | 130 | 120 | 150 | 0.009 |
| Diastolic BP, mm Hg | | | | | |
| | lowest | 78 | 70 | 84 | 0.008 |
| | low | 74 | 67 | 85 | |
| | high | 68 | 62 | 78 | |
| | highest | 74 | 65 | 88 | 0.382 |
| Respiratory Rate, breaths/minute | | | | | |
| | lowest | 18 | 18 | 20 | < 0.001 |

| | | | | | |
|---|---|---|---|---|---|
| | low | 20 | 18 | 22 | |
| | high | 20 | 18 | 24 | |
| | highest | 22 | 20 | 28 | < 0.001 |
| Oxygen Saturation, % | | | | | |
| | lowest | 94 | 92 | 96 | < 0.001 |
| | low | 91 | 90 | 95 | |
| | high | 91 | 88 | 94 | |
| | highest | 86 | 81 | 93 | < 0.001 |
| Weight, kg | | | | | |
| | lowest | 82 | 68 | 89 | 0.001 |
| | low | 89 | 77 | 100 | |
| | high | 85 | 70 | 100 | |
| | highest | 93 | 79 | 110 | 0.005 |
| QTc | | | | | |
| | lowest | 440 | 430 | 470 | 0.245 |
| | low | 450 | 420 | 470 | |
| | high | 430 | 420 | 450 | |
| | highest | 450 | 430 | 480 | 0.004 |
| Sodium, mmol/L | | | | | |
| | lowest | 140 | 130 | 140 | < 0.001 |
| | low | 140 | 130 | 140 | |
| | high | 140 | 130 | 140 | |
| | highest | 140 | 130 | 140 | 0.841 |
| Potassium, mmol/L | | | | | |
| | lowest | 3.8 | 3.6 | 4.1 | 0.126 |
| | low | 3.9 | 3.5 | 4.3 | |
| | high | 3.9 | 3.7 | 4.2 | |
| | highest | 4 | 3.6 | 4.2 | 0.607 |
| Carbon Dioxide, mmol/L | | | | | |
| | lowest | 23 | 21 | 25 | 0.097 |
| | low | 23 | 21 | 25 | |
| | high | 23 | 20 | 24 | |
| | highest | 22 | 21 | 25 | 0.516 |
| BUN, mg/dL | | | | | |
| | lowest | 13 | 9 | 18 | < 0.001 |
| | low | 15 | 11 | 22 | |
| | high | 15 | 12 | 22 | |
| | highest | 17 | 12 | 29 | 0.001 |
| Creatinine, mg/dL | | | | | |
| | lowest | 0.84 | 0.69 | 1.1 | 0.088 |
| | low | 0.89 | 0.73 | 1.3 | |
| | high | 0.82 | 0.7 | 1 | |
| | highest | 0.91 | 0.79 | 1.4 | 0.011 |

| | | | | | |
|---|---|---|---|---|---|
| Anion Gap, mmol/L | | | | | |
| | lowest | 10 | 9 | 12 | 0.008 |
| | low | 11 | 10 | 12 | |
| | high | 11 | 9 | 12 | |
| | highest | 12 | 10 | 14 | < 0.001 |
| WBC Count, 109 cells/L | | | | | |
| | lowest | 6.3 | 4.6 | 7.9 | < 0.001 |
| | low | 6.4 | 5.3 | 7.7 | |
| | high | 7.6 | 5.9 | 9.8 | |
| | highest | 8.7 | 7.1 | 11 | < 0.001 |
| Hemoglobin, g/dL | | | | | |
| | lowest | 14 | 12 | 15 | 0.07 |
| | low | 14 | 13 | 16 | |
| | high | 14 | 13 | 15 | |
| | highest | 15 | 13 | 16 | 0.141 |
| Hematocrit, % | | | | | |
| | lowest | 41 | 37 | 45 | 0.162 |
| | low | 42 | 39 | 46 | |
| | high | 42 | 39 | 45 | |
| | highest | 43 | 38 | 47 | 0.104 |
| Platelet Count, 109 cells/L | | | | | |
| | lowest | 200 | 170 | 260 | 0.563 |
| | low | 200 | 160 | 260 | |
| | high | 190 | 160 | 240 | |
| | highest | 190 | 150 | 290 | 0.925 |
| LDH, U/L | | | | | |
| | lowest | 260 | 220 | 510 | < 0.001 |
| | low | 400 | 330 | 710 | |
| | high | 570 | 330 | 900 | |
| | highest | 560 | 410 | 1000 | < 0.001 |
| D-Dimer ng/mL | | | | | |
| | lowest | 470 | 320 | 960 | < 0.001 |
| | low | 510 | 380 | 1300 | |
| | high | 790 | 590 | 1200 | |
| | highest | 1400 | 890 | 2300 | < 0.001 |
| CRP, mg/L | | | | | |
| | lowest | 31 | 14 | 56 | < 0.001 |
| | low | 68 | 36 | 160 | |
| | high | 88 | 69 | 150 | |
| | highest | 180 | 130 | 240 | < 0.001 |
| Ferritin, ng/mL | | | | | |
| | lowest | 130 | 65 | 250 | < 0.001 |
| | low | 430 | 300 | 720 | |

| | | | | | |
|---|---|---|---|---|---|
| | high | 300 | 130 | 860 | |
| | highest | 470 | 210 | 940 | < 0.001 |

#### Prediction of acute respiratory distress syndrome (ARDS)

Figures E8 through E11 compare the ROC curves in development and validation for the individual component classifiers in the ARDS test with the cutoff value of 0.5 marked for each. AUCs are also given in the legends. Tables E6 and E7 respectively summarize the categorical and continuous attributes used in prediction for each of the final risk groups of the ARDS test in development. Tables E8 and E9 give the same in the validation set. Mann-Whitney p-values are given comparing the continuous attribute ranks between the highest and lowest risk groups compared to all other risk groups. These statistics were calculated using Matlab R2020a. In the validation cohort, for race, a chi-squared test of independence was calculated with race grouped as: white, black, Hispanic, and other. The p-value was 0.519. For EGFR and gender, a Fisher exact test was used yielding p-values of 0.0189 and 0.0405 respectively. These statistics were calculated in SAS Enterprise Guide 8.2 (SAS 9.4).

**Figure E8: ROC Curve Comparison for the ARDS First Split Classifier**

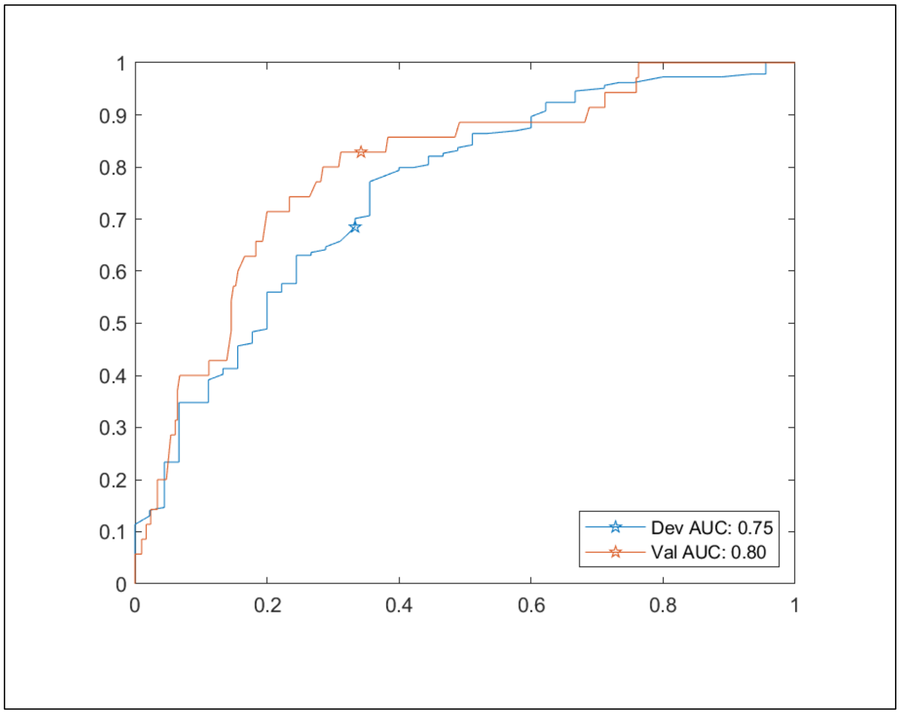

**Figure E9: ROC Curve Comparison for the ARDS Low Risk Child Classifier**

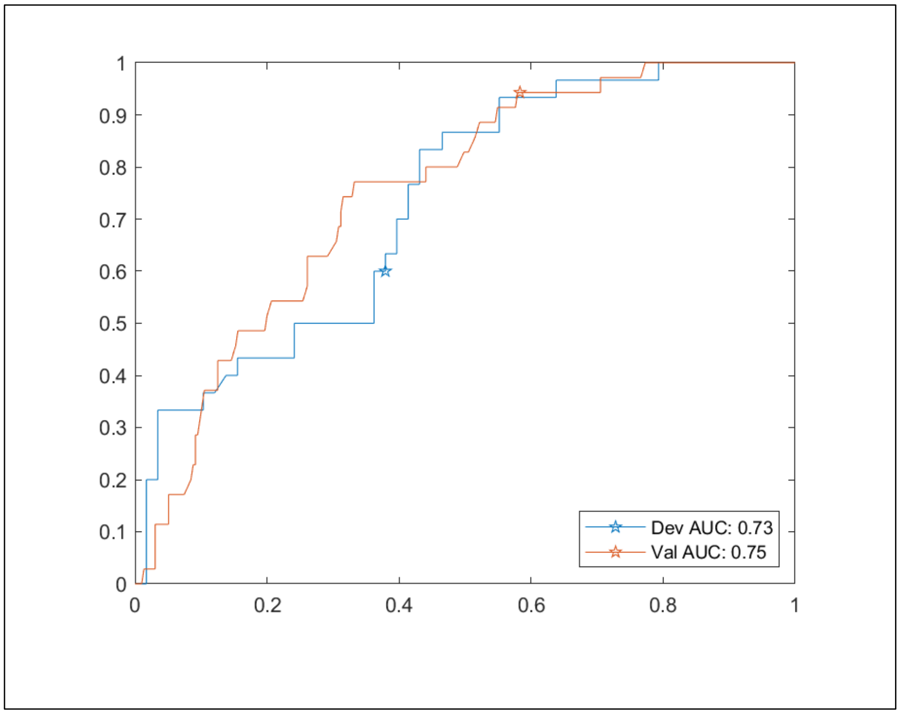


**Figure E10: ROC Curve Comparison for the ARDS High Risk Child Classifier**

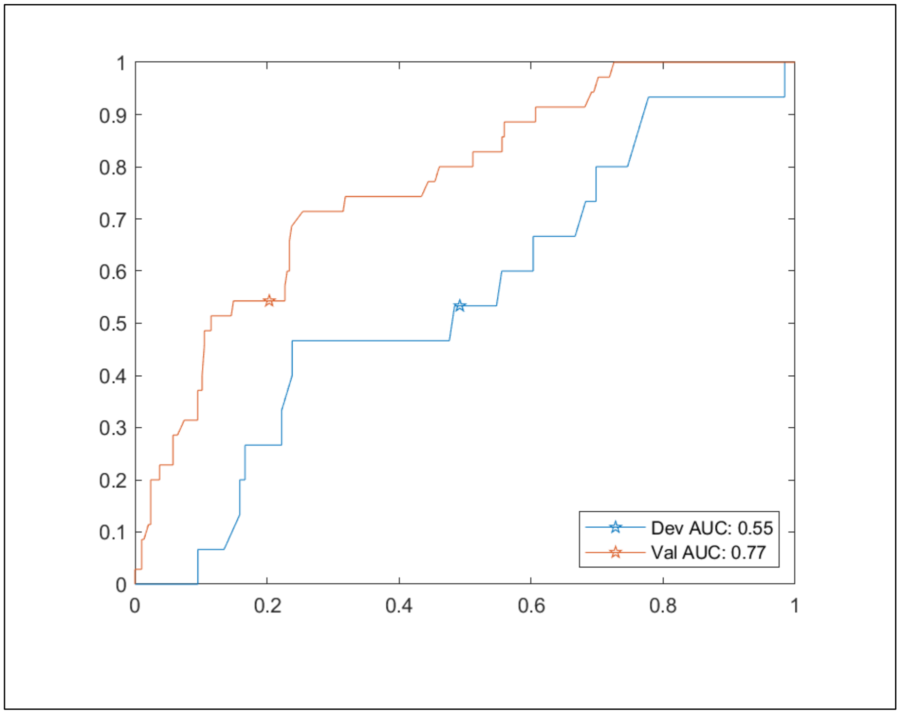

**Figure E11: ROC Curve Comparison for the ARDS Intermediate Risk Grandchild Classifier**

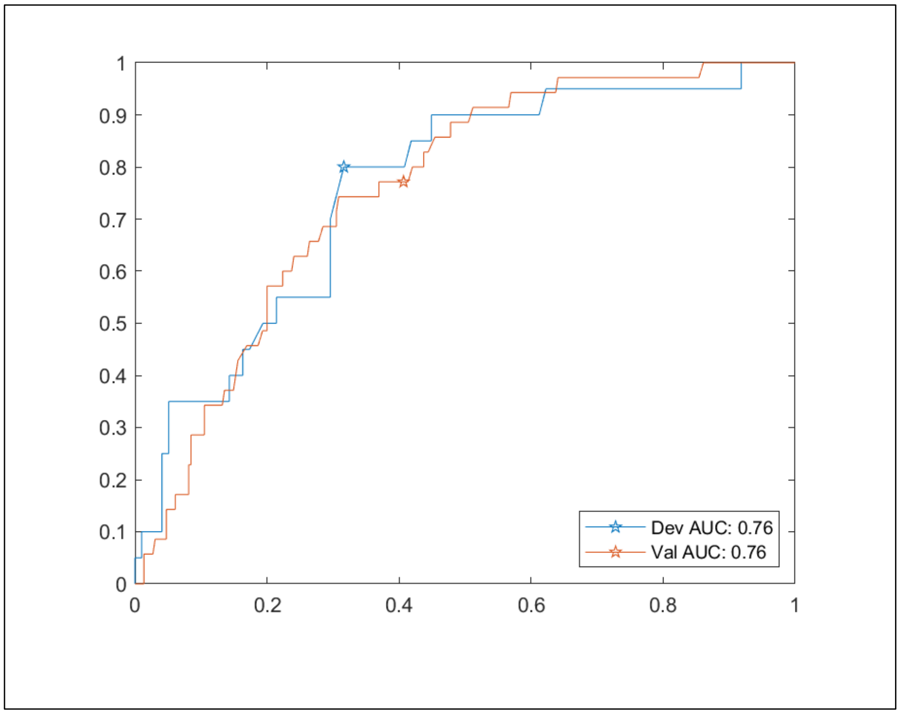


**Table E10: Summary of Categorical Attributes by ARDS Risk Group in Development**

| Attribute | Class | lowest | high | highest |
|---|---|---|---|---|
| | | n (proportion of group) | | |
| Race | | N=142 | N=47 | N=40 |
| | White | 30 (0.211) | 9 (0.191) | 2 (0.050) |
| | Black/African American | 27 (0.190) | 10 (0.213) | 15 (0.375) |
| | Hispanic/Latino | 59 (0.415) | 21 (0.447) | 14 (0.350) |
| | Other | 18 (0.127) | 7 (0.149) | 6 (0.150) |
| | Unknown | 8 (0.056) | 0 (0.000) | 3 (0.075) |
| Gender | | N=142 | N=47 | N=40 |
| | Male | 71 (0.500) | 24 (0.511) | 29 (0.725) |
| | Female | 71 (0.500) | 23 (0.489) | 11 (0.275) |
| eGFR | | N=142 | N=47 | N=40 |
| | x>=60 | 126 (0.887) | 37 (0.787) | 17 (0.425) |
| | 30>=x>60 | 8 (0.056) | 7 (0.149) | 19 (0.475) |
| | x<30 | 8 (0.056) | 3 (0.064) | 4 (0.100) |

**Table E11: Summary of Continuous Attributes by ARDS Risk Group in Development**

| Attribute | Label | Median | 25th | 75th | Mann-Whitney p |
|---|---|---|---|---|---|
| Age, years | | | | | |
| | lowest | 55 | 37 | 68 | 0.024 |
| | high | 60 | 48 | 66 | |
| | highest | 59 | 51 | 72 | 0.05 |
| Temperature, °C | | | | | |
| | lowest | 37 | 37 | 38 | 0.974 |
| | high | 37 | 37 | 38 | |
| | highest | 37 | 36 | 38 | 0.524 |
| Heart Rate, beats/minute | | | | | |
| | lowest | 98 | 84 | 110 | 0.837 |
| | high | 93 | 82 | 100 | |
| | highest | 110 | 86 | 120 | 0.078 |
| Systolic BP, mm Hg | | | | | |
| | lowest | 140 | 120 | 150 | < 0.001 |
| | high | 120 | 110 | 130 | |
| | highest | 130 | 120 | 140 | 0.604 |
| Diastolic BP, mm Hg | | | | | |
| | lowest | 77 | 68 | 85 | < 0.001 |
| | high | 68 | 59 | 75 | |
| | highest | 74 | 66 | 84 | 0.988 |
| Respiratory Rate, breaths/minute | | | | | |
| | lowest | 20 | 18 | 24 | 0.261 |
| | high | 18 | 18 | 24 | |
| | highest | 24 | 18 | 30 | 0.005 |
| Oxygen Saturation, % | | | | | |
| | lowest | 92 | 90 | 95 | < 0.001 |
| | high | 92 | 87 | 94 | |
| | highest | 85 | 76 | 90 | < 0.001 |
| Weight, kg | | | | | |
| | lowest | 80 | 72 | 97 | 0.186 |
| | high | 84 | 73 | 98 | |
| | highest | 92 | 71 | 110 | 0.198 |
| QTc | | | | | |
| | lowest | 450 | 430 | 460 | 0.067 |
| | high | 430 | 420 | 450 | |
| | highest | 440 | 420 | 460 | 0.626 |
| Sodium, mmol/L | | | | | |
| | lowest | 140 | 130 | 140 | 0.449 |
| | high | 140 | 130 | 140 | |
| | highest | 140 | 130 | 140 | 1 |

| | | | | | |
|---|---|---|---|---|---|
| Potassium, mmol/L | | | | | |
| | lowest | 3.7 | 3.4 | 3.9 | 0.039 |
| | high | 3.8 | 3.6 | 4.1 | |
| | highest | 3.8 | 3.4 | 4.2 | 0.203 |
| Carbon Dioxide, mmol/L | | | | | |
| | lowest | 23 | 22 | 25 | < 0.001 |
| | high | 22 | 20 | 23 | |
| | highest | 22 | 19 | 24 | 0.022 |
| BUN, mg/dL | | | | | |
| | lowest | 12 | 9 | 16 | < 0.001 |
| | high | 15 | 11 | 24 | |
| | highest | 27 | 15 | 38 | < 0.001 |
| Creatinine, mg/dL | | | | | |
| | lowest | 0.82 | 0.64 | 1.1 | < 0.001 |
| | high | 0.99 | 0.74 | 1.2 | |
| | highest | 1.3 | 0.98 | 1.7 | < 0.001 |
| Anion Gap, mmol/L | | | | | |
| | lowest | 11 | 10 | 13 | < 0.001 |
| | high | 12 | 10 | 13 | |
| | highest | 13 | 12 | 16 | < 0.001 |
| WBC Count, 109 cells/L | | | | | |
| | lowest | 6 | 4.7 | 7.6 | < 0.001 |
| | high | 7.3 | 6.1 | 10 | |
| | highest | 9.8 | 7.2 | 15 | < 0.001 |
| Hemoglobin, g/dL | | | | | |
| | lowest | 15 | 14 | 16 | 0.102 |
| | high | 14 | 13 | 15 | |
| | highest | 15 | 14 | 16 | 0.035 |
| Hematocrit, % | | | | | |
| | lowest | 44 | 41 | 47 | 0.14 |
| | high | 40 | 38 | 44 | |
| | highest | 45 | 41 | 49 | 0.014 |
| Platelet Count, 109 cells/L | | | | | |
| | lowest | 210 | 170 | 250 | 0.899 |
| | high | 220 | 150 | 270 | |
| | highest | 200 | 150 | 300 | 0.968 |
| LDH, U/L | | | | | |
| | lowest | 290 | 230 | 340 | < 0.001 |
| | high | 320 | 250 | 430 | |
| | highest | 500 | 420 | 640 | < 0.001 |
| D-Dimer ng/mL | | | | | |
| | lowest | 600 | 420 | 910 | < 0.001 |
| | high | 1000 | 870 | 2000 | |

| | | | | | |
|---|---|---|---|---|---|
| | highest | 1500 | 1100 | 2700 | < 0.001 |
| CRP, mg/L | | | | | |
| | lowest | 57 | 27 | 94 | < 0.001 |
| | high | 130 | 89 | 170 | |
| | highest | 220 | 130 | 300 | < 0.001 |
| Ferritin, ng/mL | | | | | |
| | lowest | 300 | 140 | 530 | < 0.001 |
| | high | 430 | 180 | 900 | |
| | highest | 850 | 400 | 1300 | < 0.001 |

**Table E12: Summary of Categorical Attributes by ARDS Risk Group in Validation**

| Attribute | Class | lowest | high | highest |
|---|---|---|---|---|
| | | n (proportion of group) | | |
| Race | | | | |
| | White | 68 (0.358) | 28 (0.384) | 17 (0.254) |
| | Black/African American | 15 (0.079) | 5 (0.068) | 3 (0.045) |
| | Hispanic/Latino | 89 (0.468) | 34 (0.466) | 41 (0.612) |
| | Other | 17 (0.089) | 5 (0.068) | 3 (0.045) |
| | Unknown | 1 (0.005) | 1 (0.014) | 3 (0.045) |
| Gender | | | | |
| | Male | 91 (0.479) | 37 (0.507) | 44 (0.657) |
| | Female | 99 (0.521) | 36 (0.493) | 23 (0.343) |
| eGFR | | | | |
| | x>=60 | 146 (0.768) | 54 (0.740) | 44 (0.657) |
| | 30>=x>60 | 37 (0.195) | 12 (0.164) | 12 (0.179) |
| | x<30 | 7 (0.037) | 7 (0.096) | 11 (0.164) |

**Table E13: Summary of Continuous Attributes by ARDS Risk Group in Validation**

| Attribute | Label | Median | 25th | 75th | Mann-Whitney p |
|---|---|---|---|---|---|
| Age, years | | | | | |
| | lowest | 52 | 40 | 68 | < 0.001 |
| | high | 60 | 50 | 73 | |
| | highest | 60 | 53 | 73 | 0.009 |
| Temperature, °C | | | | | |
| | lowest | 37 | 37 | 38 | 0.206 |
| | high | 37 | 37 | 37 | |
| | highest | 37 | 37 | 38 | 0.288 |
| Heart Rate, beats/minute | | | | | |
| | lowest | 98 | 86 | 110 | 0.386 |
| | high | 92 | 81 | 110 | |
| | highest | 100 | 89 | 110 | 0.079 |
| Systolic BP, mm Hg | | | | | |
| | lowest | 130 | 120 | 140 | 0.228 |
| | high | 120 | 110 | 130 | |
| | highest | 140 | 120 | 150 | 0.013 |
| Diastolic BP, mm Hg | | | | | |
| | lowest | 77 | 67 | 87 | < 0.001 |
| | high | 66 | 61 | 73 | |
| | highest | 74 | 64 | 83 | 0.741 |
| Respiratory Rate, breaths/minute | | | | | |
| | lowest | 19 | 18 | 22 | < 0.001 |
| | high | 20 | 18 | 24 | |
| | highest | 22 | 20 | 26 | < 0.001 |
| Oxygen Saturation, % | | | | | |
| | lowest | 93 | 90 | 95 | < 0.001 |
| | high | 92 | 87 | 95 | |
| | highest | 85 | 79 | 91 | < 0.001 |
| Weight, kg | | | | | |
| | lowest | 86 | 74 | 100 | 0.757 |
| | high | 82 | 69 | 99 | |
| | highest | 86 | 77 | 110 | 0.184 |
| QTc | | | | | |
| | lowest | 440 | 430 | 460 | 0.664 |
| | high | 440 | 420 | 470 | |
| | highest | 450 | 430 | 470 | 0.015 |
| Sodium, mmol/L | | | | | |
| | lowest | 140 | 130 | 140 | < 0.001 |
| | high | 140 | 130 | 140 | |
| | highest | 130 | 130 | 140 | 0.007 |

| | | | | | |
|---|---|---|---|---|---|
| Potassium, mmol/L | | | | | |
| | lowest | 3.8 | 3.6 | 4.2 | 0.167 |
| | high | 3.9 | 3.6 | 4.2 | |
| | highest | 4 | 3.7 | 4.3 | 0.116 |
| Carbon Dioxide, mmol/L | | | | | |
| | lowest | 23 | 21 | 25 | 0.345 |
| | high | 23 | 20 | 24 | |
| | highest | 22 | 21 | 25 | 0.696 |
| BUN, mg/dL | | | | | |
| | lowest | 13 | 9 | 19 | < 0.001 |
| | high | 14 | 12 | 23 | |
| | highest | 21 | 13 | 33 | < 0.001 |
| Creatinine, mg/dL | | | | | |
| | lowest | 0.85 | 0.72 | 1.1 | 0.071 |
| | high | 0.84 | 0.71 | 1.2 | |
| | highest | 0.93 | 0.8 | 1.4 | 0.006 |
| Anion Gap, mmol/L | | | | | |
| | lowest | 11 | 9 | 13 | 0.736 |
| | high | 10 | 8.8 | 12 | |
| | highest | 12 | 9.3 | 14 | 0.014 |
| WBC Count, 109 cells/L | | | | | |
| | lowest | 6.3 | 5 | 7.8 | < 0.001 |
| | high | 7.8 | 5.8 | 10 | |
| | highest | 9.2 | 7.3 | 12 | < 0.001 |
| Hemoglobin, g/dL | | | | | |
| | lowest | 14 | 13 | 15 | 0.488 |
| | high | 14 | 13 | 15 | |
| | highest | 15 | 13 | 16 | 0.203 |
| Hematocrit, % | | | | | |
| | lowest | 42 | 39 | 45 | 0.436 |
| | high | 40 | 38 | 44 | |
| | highest | 43 | 38 | 47 | 0.242 |
| Platelet Count, 109 cells/L | | | | | |
| | lowest | 200 | 160 | 250 | 0.638 |
| | high | 190 | 140 | 250 | |
| | highest | 210 | 160 | 300 | 0.259 |
| LDH, U/L | | | | | |
| | lowest | 340 | 250 | 610 | < 0.001 |
| | high | 500 | 330 | 770 | |
| | highest | 810 | 500 | 1000 | < 0.001 |
| D-Dimer ng/mL | | | | | |
| | lowest | 490 | 330 | 800 | < 0.001 |
| | high | 1300 | 790 | 2500 | |

| | | | | | |
|---|---|---|---|---|---|
| | highest | 1300 | 890 | 1700 | < 0.001 |
| CRP, mg/L | | | | | |
| | lowest | 49 | 22 | 85 | < 0.001 |
| | high | 130 | 69 | 190 | |
| | highest | 180 | 140 | 220 | < 0.001 |
| Ferritin, ng/mL | | | | | |
| | lowest | 240 | 94 | 510 | < 0.001 |
| | high | 340 | 170 | 780 | |
| | highest | 450 | 270 | 940 | < 0.001 |

#### Prediction of Intubation

Figures E12 through E13 compare the ROC curves in development and validation for the individual component classifiers in the intubation test with the cutoff value of 0.5 marked for each. AUCs are also given in the legends. Tables E10 and E11 respectively summarize the categorical and continuous attributes used in prediction for each of the final risk groups of the intubation test in development. Tables E12 and E13 give the same in the validation set. Mann-Whitney p-values are given comparing the continuous attribute ranks between the highest and lowest risk groups compared to all other risk groups. These statistics were calculated using Matlab R2020a. In the validation cohort, for race, a chi-squared test of independence was calculated with race grouped as: white, black, Hispanic, and other. The p-value was less than 0.0001. For EGFR and gender, a Fisher exact test was used yielding p-values of 0.0005 and 0.0873 respectively. These statistics were calculated using SAS Enterprise Guide 8.2 (SAS 9.4).

**Figure E12: ROC Curve Comparison for the Intubation First Split Classifier**

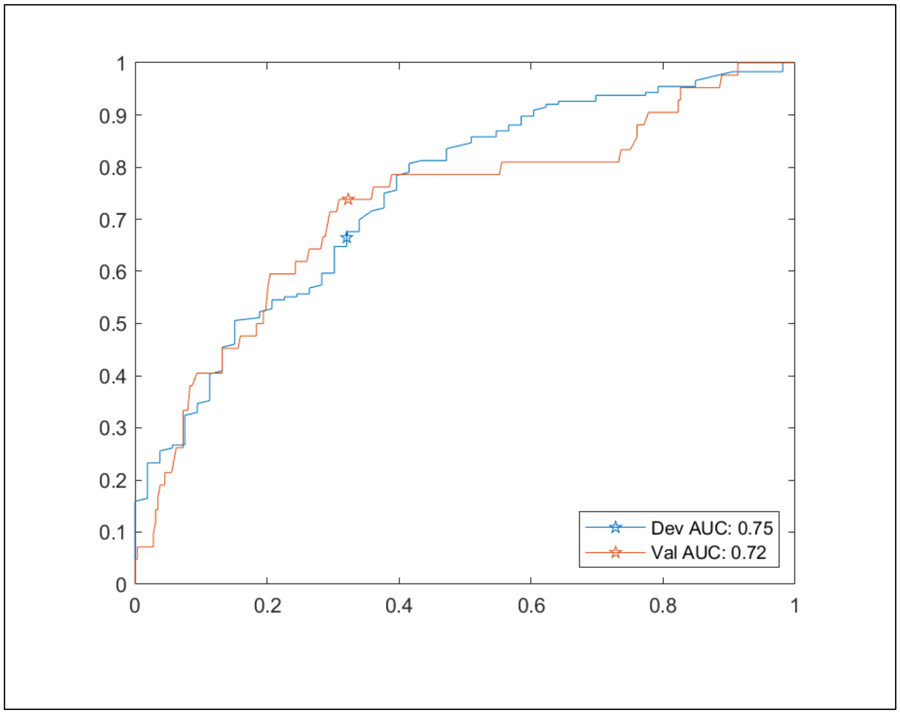

**Figure E13: ROC Curve Comparison for the Intubation Low Risk Child Classifier**

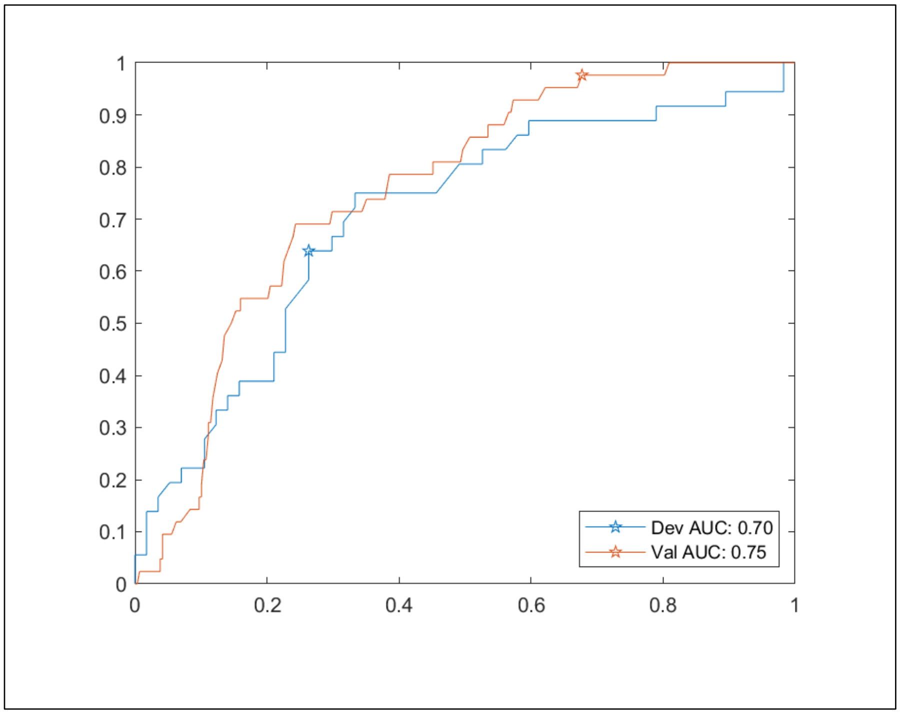


**Figure E14: ROC Curve Comparison for the Intubation High Risk Child Classifier**

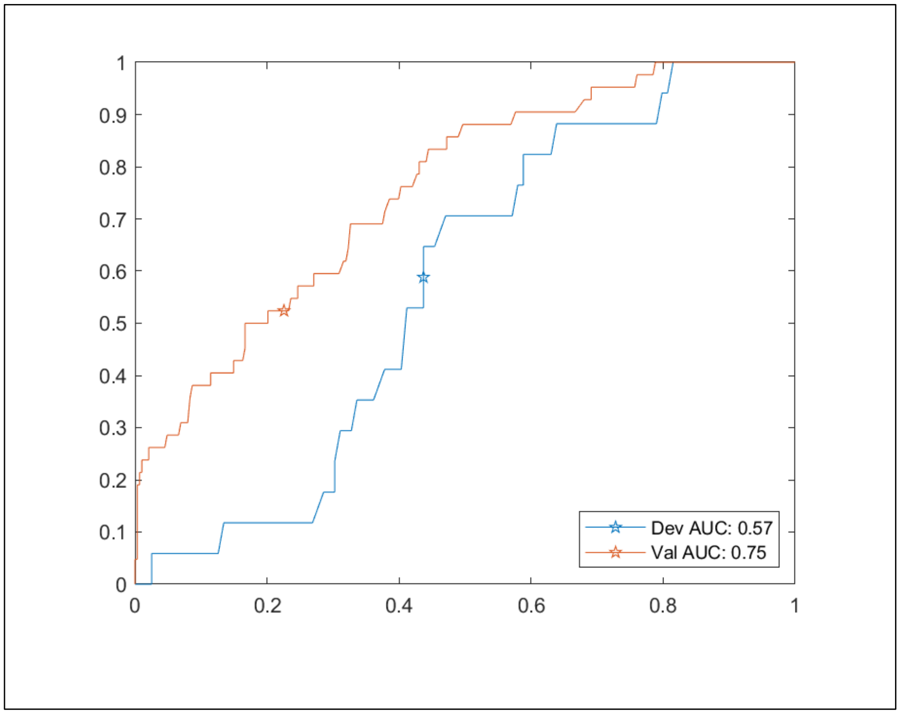

**Table E14: Summary of Categorical Attributes by Intubation Risk Group in Development**

| Attribute | Class | lowest | low | intermediate | highest |
|---|---|---|---|---|---|
| | | n (proportion of group) | | | |
| Race | | N=74 | N=62 | N=55 | N=38 |
| | White | 19 (0.257) | 20 (0.323) | 1 (0.018) | 1 (0.026) |
| | Black/African American | 9 (0.122) | 6 (0.097) | 22 (0.400) | 15 (0.395) |
| | Hispanic/Latino | 34 (0.459) | 29 (0.468) | 20 (0.364) | 11 (0.289) |
| | Other | 7 (0.095) | 4 (0.065) | 12 (0.218) | 8 (0.211) |
| | Unknown | 5 (0.068) | 3 (0.048) | 0 (0.000) | 3 (0.079) |
| Gender | | N=74 | N=62 | N=55 | N=38 |
| | Male | 38 (0.514) | 26 (0.419) | 33 (0.600) | 27 (0.711) |
| | Female | 36 (0.486) | 36 (0.581) | 22 (0.400) | 11 (0.289) |
| eGFR | | N=74 | N=62 | N=55 | N=38 |
| | x>=60 | 67 (0.905) | 54 (0.871) | 46 (0.836) | 13 (0.342) |
| | 30>=x>60 | 3 (0.041) | 6 (0.097) | 4 (0.073) | 21 (0.553) |
| | x<30 | 4 (0.054) | 2 (0.032) | 5 (0.091) | 4 (0.105) |

**Table E15: Summary of Continuous Attributes by Intubation Risk Group in Development**

| Attribute | Label | Median | 25$^{th}$ Percentile | 75$^{th}$ Percentile | Mann-Whitney p |
|---|---|---|---|---|---|
| Age, years | | | | | |
| | lowest | 51 | 36 | 64 | 0.004 |
| | low | 55 | 34 | 68 | |
| | intermediate | 62 | 52 | 69 | |
| | highest | 60 | 52 | 72 | 0.015 |
| Temperature, °C | | | | | |
| | lowest | 37 | 36 | 37 | 0.05 |
| | low | 37 | 37 | 38 | |
| | intermediate | 37 | 37 | 38 | |
| | highest | 37 | 36 | 37 | 0.098 |
| Heart Rate, beats/minute | | | | | |
| | lowest | 97 | 81 | 110 | 0.154 |
| | low | 100 | 89 | 120 | |
| | intermediate | 88 | 83 | 110 | |
| | highest | 110 | 88 | 120 | 0.021 |
| Systolic BP, mm Hg | | | | | |
| | lowest | 130 | 120 | 150 | 0.297 |
| | low | 140 | 120 | 150 | |
| | intermediate | 130 | 120 | 150 | |

| | | | | | |
|---|---|---|---|---|---|
| | highest | 130 | 120 | 140 | 0.431 |
| Diastolic BP, mm Hg | | | | | |
| | lowest | 74 | 64 | 87 | 0.805 |
| | low | 74 | 68 | 82 | |
| | intermediate | 74 | 66 | 80 | |
| | highest | 74 | 65 | 85 | 0.614 |
| Respiratory Rate, breaths/minute | | | | | |
| | lowest | 20 | 18 | 23 | 0.169 |
| | low | 20 | 18 | 24 | |
| | intermediate | 20 | 18 | 25 | |
| | highest | 24 | 18 | 30 | 0.007 |
| Oxygen Saturation, % | | | | | |
| | lowest | 93 | 90 | 95 | < 0.001 |
| | low | 92 | 88 | 95 | |
| | intermediate | 92 | 88 | 94 | |
| | highest | 85 | 75 | 87 | < 0.001 |
| Weight, kg | | | | | |
| | lowest | 77 | 72 | 91 | 0.065 |
| | low | 80 | 73 | 91 | |
| | intermediate | 89 | 70 | 110 | |
| | highest | 93 | 71 | 110 | 0.102 |
| QTc | | | | | |
| | lowest | 440 | 420 | 460 | 0.354 |
| | low | 440 | 420 | 450 | |
| | intermediate | 450 | 430 | 460 | |
| | highest | 440 | 420 | 470 | 0.583 |
| Sodium, mmol/L | | | | | |
| | lowest | 140 | 130 | 140 | 0.319 |
| | low | 140 | 130 | 140 | |
| | intermediate | 140 | 130 | 140 | |
| | highest | 140 | 130 | 140 | 0.985 |
| Potassium, mmol/L | | | | | |
| | lowest | 3.7 | 3.5 | 4 | 0.794 |
| | low | 3.8 | 3.4 | 3.9 | |
| | intermediate | 3.7 | 3.4 | 3.9 | |
| | highest | 3.8 | 3.5 | 4.2 | 0.082 |
| Carbon Dioxide, mmol/L | | | | | |
| | lowest | 24 | 22 | 25 | < 0.001 |
| | low | 22 | 21 | 24 | |
| | intermediate | 23 | 22 | 24 | |
| | highest | 21 | 19 | 24 | 0.013 |
| BUN, mg/dL | | | | | |

| | | | | | |
|---|---|---|---|---|---|
| | lowest | 11 | 7 | 15 | < 0.001 |
| | low | 13 | 10 | 18 | |
| | intermediate | 14 | 11 | 19 | |
| | highest | 27 | 18 | 38 | < 0.001 |
| Creatinine, mg/dL | | | | | |
| | lowest | 0.79 | 0.63 | 1 | < 0.001 |
| | low | 0.81 | 0.68 | 1.1 | |
| | intermediate | 0.99 | 0.72 | 1.2 | |
| | highest | 1.4 | 1.1 | 1.8 | < 0.001 |
| Anion Gap, mmol/L | | | | | |
| | lowest | 11 | 9 | 12 | < 0.001 |
| | low | 12 | 11 | 14 | |
| | intermediate | 12 | 10 | 13 | |
| | highest | 14 | 12 | 16 | < 0.001 |
| WBC Count, 109 cells/L | | | | | |
| | lowest | 5.7 | 4.2 | 6.7 | < 0.001 |
| | low | 7.3 | 5.6 | 8.9 | |
| | intermediate | 7.1 | 5.6 | 8.4 | |
| | highest | 9.6 | 7.1 | 15 | < 0.001 |
| Hemoglobin, g/dL | | | | | |
| | lowest | 15 | 14 | 16 | 0.037 |
| | low | 14 | 13 | 16 | |
| | intermediate | 14 | 13 | 15 | |
| | highest | 15 | 14 | 16 | 0.244 |
| Hematocrit, % | | | | | |
| | lowest | 44 | 41 | 47 | 0.07 |
| | low | 42 | 39 | 46 | |
| | intermediate | 43 | 39 | 45 | |
| | highest | 45 | 40 | 49 | 0.117 |
| Platelet Count, 109 cells/L | | | | | |
| | lowest | 190 | 160 | 250 | 0.105 |
| | low | 230 | 190 | 280 | |
| | intermediate | 220 | 160 | 250 | |
| | highest | 180 | 140 | 300 | 0.398 |
| LDH, U/L | | | | | |
| | lowest | 270 | 230 | 330 | < 0.001 |
| | low | 310 | 250 | 380 | |
| | intermediate | 330 | 280 | 450 | |
| | highest | 490 | 390 | 650 | < 0.001 |
| D-Dimer ng/mL | | | | | |
| | lowest | 580 | 390 | 910 | < 0.001 |
| | low | 780 | 550 | 1500 | |
| | intermediate | 940 | 650 | 1500 | |

| | | | | | |
|---|---|---|---|---|---|
| | highest | 1400 | 920 | 2800 | < 0.001 |
| CRP, mg/L | | | | | |
| | lowest | 41 | 19 | 66 | < 0.001 |
| | low | 98 | 66 | 160 | |
| | intermediate | 110 | 58 | 160 | |
| | highest | 200 | 130 | 310 | < 0.001 |
| Ferritin, ng/mL | | | | | |
| | lowest | 340 | 160 | 530 | 0.135 |
| | low | 230 | 130 | 440 | |
| | intermediate | 520 | 210 | 1000 | |
| | highest | 640 | 360 | 1200 | < 0.001 |

**Table E16: Summary of Categorical Attributes by Intubation Risk Group in Validation**

| Attribute | Class | lowest | low | intermediate | highest |
|---|---|---|---|---|---|
| | | n (proportion of group) | | | |
| Race | | | | | |
| | White | 36 (0.419) | 60 (0.500) | 9 (0.134) | 8 (0.140) |
| | Black/African American | 4 (0.047) | 3 (0.025) | 10 (0.149) | 6 (0.105) |
| | Hispanic/Latino | 35 (0.407) | 54 (0.450) | 38 (0.567) | 37 (0.649) |
| | Other | 11 (0.128) | 2 (0.017) | 9 (0.134) | 3 (0.053) |
| | Unknown | 0 (0.000) | 1 (0.008) | 1 (0.015) | 3 (0.053) |
| Gender | | | | | |
| | Male | 44 (0.512) | 53 (0.442) | 40 (0.597) | 35 (0.614) |
| | Female | 42 (0.488) | 67 (0.558) | 27 (0.403) | 22 (0.386) |
| eGFR | | | | | |
| | x>=60 | 66 (0.767) | 95 (0.792) | 53 (0.791) | 30 (0.526) |
| | 30>=x>60 | 15 (0.174) | 19 (0.158) | 13 (0.194) | 14 (0.246) |
| | x<30 | 5 (0.058) | 6 (0.050) | 1 (0.015) | 13 (0.228) |

**Table E17: Summary of Continuous Attributes by Intubation Risk Group in Validation**

| Attribute | Label | Median | 25th | 75th | Mann-Whitney p |
|---|---|---|---|---|---|
| Age, years | | | | | |
| | lowest | 56 | 38 | 68 | 0.027 |
| | low | 58 | 38 | 72 | |
| | intermediate | 54 | 50 | 64 | |
| | highest | 61 | 55 | 72 | 0.004 |
| Temperature, °C | | | | | |
| | lowest | 37 | 36 | 37 | 0.002 |

| | | | | | |
|---|---|---|---|---|---|
| | low | 37 | 37 | 38 | |
| | intermediate | 37 | 37 | 38 | |
| | highest | 37 | 37 | 38 | 0.763 |
| Heart Rate, beats/minute | | | | | |
| | lowest | 94 | 80 | 100 | 0.004 |
| | low | 100 | 86 | 120 | |
| | intermediate | 96 | 85 | 110 | |
| | highest | 100 | 91 | 110 | 0.067 |
| Systolic BP, mm Hg | | | | | |
| | lowest | 130 | 120 | 140 | 0.488 |
| | low | 130 | 120 | 140 | |
| | intermediate | 130 | 120 | 140 | |
| | highest | 130 | 120 | 150 | 0.071 |
| Diastolic BP, mm Hg | | | | | |
| | lowest | 77 | 66 | 85 | 0.284 |
| | low | 72 | 65 | 82 | |
| | intermediate | 74 | 65 | 83 | |
| | highest | 73 | 61 | 85 | 0.565 |
| Respiratory Rate, breaths/minute | | | | | |
| | lowest | 18 | 18 | 20 | < 0.001 |
| | low | 20 | 18 | 24 | |
| | intermediate | 20 | 18 | 24 | |
| | highest | 22 | 20 | 25 | < 0.001 |
| Oxygen Saturation, % | | | | | |
| | lowest | 93 | 91 | 95 | 0.001 |
| | low | 92 | 89 | 95 | |
| | intermediate | 91 | 88 | 94 | |
| | highest | 88 | 82 | 93 | < 0.001 |
| Weight, kg | | | | | |
| | lowest | 82 | 71 | 100 | 0.186 |
| | low | 82 | 73 | 94 | |
| | intermediate | 98 | 82 | 110 | |
| | highest | 91 | 76 | 110 | 0.168 |
| QTc | | | | | |
| | lowest | 440 | 430 | 460 | 0.497 |
| | low | 440 | 420 | 470 | |
| | intermediate | 440 | 420 | 460 | |
| | highest | 450 | 440 | 480 | 0.002 |
| Sodium, mmol/L | | | | | |
| | lowest | 140 | 130 | 140 | 0.083 |
| | low | 140 | 130 | 140 | |
| | intermediate | 140 | 130 | 140 | |

| | | | | | |
|---|---|---|---|---|---|
| | highest | 140 | 130 | 140 | 0.308 |
| Potassium, mmol/L | | | | | |
| | lowest | 3.9 | 3.6 | 4.2 | 0.719 |
| | low | 3.8 | 3.5 | 4.1 | |
| | intermediate | 3.9 | 3.6 | 4.1 | |
| | highest | 4 | 3.7 | 4.4 | 0.002 |
| Carbon Dioxide, mmol/L | | | | | |
| | lowest | 23 | 22 | 25 | 0.03 |
| | low | 23 | 21 | 25 | |
| | intermediate | 23 | 21 | 25 | |
| | highest | 21 | 19 | 24 | 0.004 |
| BUN, mg/dL | | | | | |
| | lowest | 13 | 9 | 17 | 0.002 |
| | low | 13 | 10 | 20 | |
| | intermediate | 15 | 11 | 20 | |
| | highest | 25 | 16 | 45 | < 0.001 |
| Creatinine, mg/dL | | | | | |
| | lowest | 0.87 | 0.71 | 1.1 | 0.736 |
| | low | 0.83 | 0.69 | 0.98 | |
| | intermediate | 0.87 | 0.73 | 1 | |
| | highest | 1.2 | 0.82 | 2.1 | < 0.001 |
| Anion Gap, mmol/L | | | | | |
| | lowest | 11 | 9 | 12 | 0.124 |
| | low | 11 | 9 | 13 | |
| | intermediate | 10 | 9 | 12 | |
| | highest | 13 | 10 | 15 | < 0.001 |
| WBC Count, 109 cells/L | | | | | |
| | lowest | 5.6 | 4.4 | 6.8 | < 0.001 |
| | low | 7.3 | 5.7 | 9.6 | |
| | intermediate | 7.3 | 5.9 | 9.2 | |
| | highest | 9 | 7.5 | 13 | < 0.001 |
| Hemoglobin, g/dL | | | | | |
| | lowest | 15 | 13 | 15 | 0.381 |
| | low | 14 | 13 | 15 | |
| | intermediate | 14 | 13 | 15 | |
| | highest | 14 | 12 | 16 | 0.608 |
| Hematocrit, % | | | | | |
| | lowest | 43 | 38 | 46 | 0.394 |
| | low | 41 | 39 | 45 | |
| | intermediate | 41 | 38 | 45 | |
| | highest | 43 | 38 | 48 | 0.383 |
| Platelet Count, 109 cells/L | | | | | |
| | lowest | 190 | 150 | 230 | 0.136 |

| | | | | | |
|---|---|---|---|---|---|
| | low | 200 | 160 | 260 | |
| | intermediate | 210 | 170 | 250 | |
| | highest | 210 | 150 | 290 | 0.257 |
| LDH, U/L | | | | | |
| | lowest | 270 | 220 | 380 | < 0.001 |
| | low | 490 | 330 | 700 | |
| | intermediate | 520 | 360 | 830 | |
| | highest | 790 | 440 | 1100 | < 0.001 |
| D-Dimer ng/mL | | | | | |
| | lowest | 510 | 320 | 1000 | 0.003 |
| | low | 680 | 410 | 1300 | |
| | intermediate | 650 | 380 | 1300 | |
| | highest | 1400 | 920 | 2500 | < 0.001 |
| CRP, mg/L | | | | | |
| | lowest | 32 | 15 | 61 | < 0.001 |
| | low | 85 | 45 | 160 | |
| | intermediate | 120 | 64 | 180 | |
| | highest | 170 | 130 | 220 | < 0.001 |
| Ferritin, ng/mL | | | | | |
| | lowest | 320 | 100 | 660 | 0.792 |
| | low | 230 | 110 | 490 | |
| | intermediate | 310 | 170 | 590 | |
| | highest | 470 | 270 | 910 | < 0.001 |

#### Predicting Any Complication

Figures E15 and E16 give the ROC curves in development for the individual component classifiers in the any complication test. Figure E17 schematically shows how the patients are split into the four final groups.

Sixty-eight patients out of the 229 in the development set developed a complication. The initial binary classifier separated these patients into a lower and higher risk group. The lower risk group had 127 patients of which 26 developed a complication. The higher risk group had 135 patients of which 22developed a complication. Both groups were split again into higher and lower risk groups using the high and low risk child classifiers.

Patients from the lower risk group that were classified as lower risk by the low risk child classifier were assigned to the lowest risk final group consisting of 73 patients of which 9 developed a complication. The remaining lower risk group patients were assigned to the final low risk group consisting of 60 patients of which 13 developed a complication.

Patients from the higher risk group who were classified as higher risk by the high risk child classifier were assigned to the highest risk final group, consisting of 40 patients of whom 23 developed a complication. The remaining higher risk group patients were assigned to the final high risk group, consisting of 54 patients of whom 23 developed a complication.

Tables E14 and E15 summarize selected attributes by final group classification.

**Figure E15: ROC Curve for the First Split Classifier Predicting Any Complication**

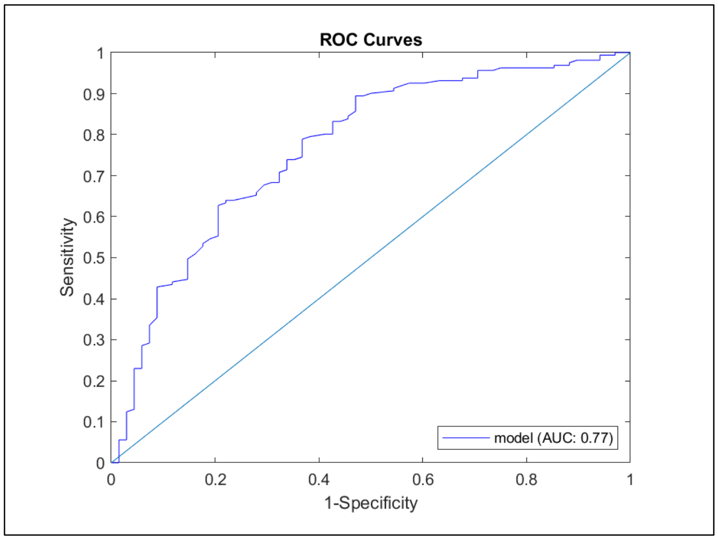


**Figure E16: ROC Curves for the Low Risk and High Risk Child Classifiers Predicting Any Complication**

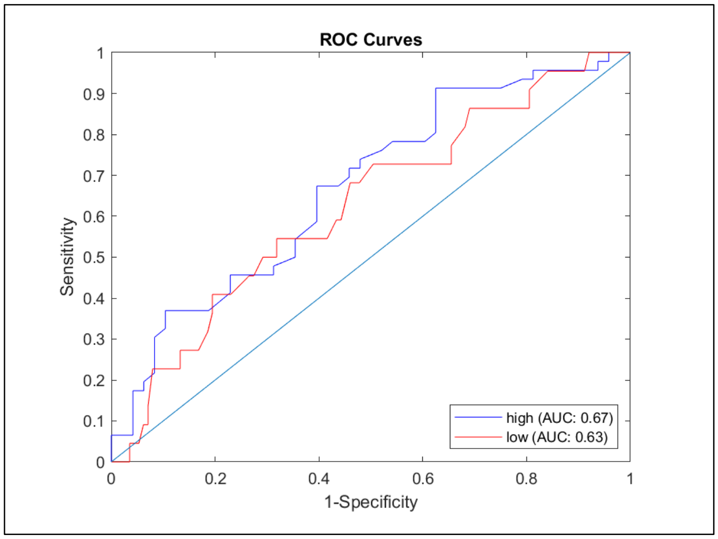

**Figure E17: Results Schematic for the Hierarchical Classifier Predicting Any Complication**

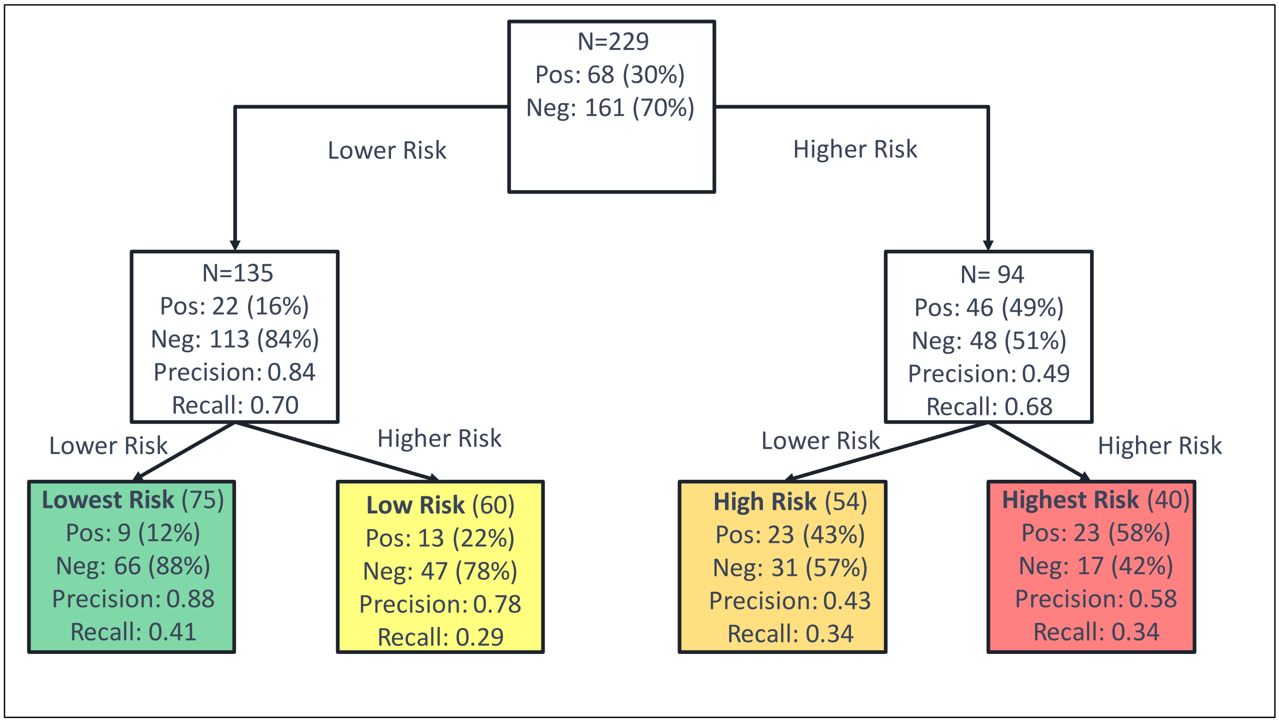


**Table E18: Summary of Categorical Attributes by Any Complication Risk Group in Development**

| Attribute | Class | lowest | low | high | highest |
|---|---|---|---|---|---|
| | | n (proportion of group) | | | |
| Race | | N=75 | N=60 | N=54 | N=40 |
| | White | 15 (0.200) | 17 (0.283) | 6 (0.111) | 3 (0.075) |
| | Black/African American | 9 (0.120) | 6 (0.100) | 22 (0.407) | 15 (0.375) |
| | Hispanic/Latino | 35 (0.467) | 30 (0.500) | 16 (0.296) | 13 (0.325) |
| | Other | 11 (0.147) | 6 (0.100) | 8 (0.148) | 6 (0.150) |
| | Unknown | 5 (0.067) | 1 (0.017) | 2 (0.037) | 3 (0.075) |
| Gender | | N=75 | N=60 | N=54 | N=40 |
| | Male | 35 (0.467) | 26 (0.433) | 35 (0.648) | 28 (0.700) |
| | Female | 40 (0.533) | 34 (0.567) | 19 (0.352) | 12 (0.300) |
| eGFR | | N=75 | N=60 | N=54 | N=40 |
| | x>=60 | 69 (0.920) | 56 (0.933) | 39 (0.722) | 16 (0.400) |
| | 30>=x>60 | 2 (0.027) | 4 (0.067) | 10 (0.185) | 18 (0.450) |
| | x<30 | 4 (0.053) | 0 (0.000) | 5 (0.093) | 6 (0.150) |

**Table E19: Summary of Continuous Attributes by Any Complication Risk Group in Development**

| Attribute | Label | Median | 25th | 75th | Mann-Whitney p |
|---|---|---|---|---|---|
| Age, years | | | | | |
| | lowest | 48 | 36 | 61 | < 0.001 |
| | low | 54 | 36 | 62 | |
| | high | 65 | 55 | 75 | |
| | highest | 63 | 52 | 73 | 0.004 |
| Temperature, Â°C | | | | | |
| | lowest | 37 | 36 | 37 | 0.013 |
| | low | 37 | 37 | 38 | |
| | high | 37 | 37 | 38 | |
| | highest | 37 | 36 | 38 | 0.565 |
| Heart Rate, beats/minute | | | | | |
| | lowest | 96 | 82 | 110 | 0.136 |
| | low | 100 | 89 | 120 | |
| | high | 95 | 84 | 110 | |
| | highest | 110 | 86 | 120 | 0.038 |
| Systolic BP, mm Hg | | | | | |
| | lowest | 130 | 110 | 150 | 0.366 |
| | low | 130 | 120 | 150 | |
| | high | 130 | 120 | 150 | |
| | highest | 130 | 120 | 140 | 0.936 |
| Diastolic BP, mm Hg | | | | | |
| | lowest | 75 | 65 | 86 | 0.284 |
| | low | 74 | 66 | 82 | |
| | high | 74 | 63 | 80 | |
| | highest | 74 | 66 | 85 | 0.927 |
| Respiratory Rate, breaths/minute | | | | | |
| | lowest | 20 | 18 | 23 | 0.486 |
| | low | 20 | 18 | 24 | |
| | high | 20 | 18 | 24 | |
| | highest | 24 | 19 | 30 | < 0.001 |
| Oxygen Saturation, % | | | | | |
| | lowest | 93 | 90 | 95 | 0.002 |
| | low | 92 | 89 | 95 | |
| | high | 91 | 87 | 94 | |
| | highest | 85 | 76 | 91 | < 0.001 |
| Weight, kg | | | | | |
| | lowest | 78 | 72 | 89 | 0.059 |

| | | | | | |
|---|---|---|---|---|---|
| | low | 79 | 71 | 90 | |
| | high | 99 | 82 | 110 | |
| | highest | 93 | 70 | 110 | 0.236 |
| QTc | | | | | |
| | lowest | 430 | 410 | 460 | 0.048 |
| | low | 440 | 420 | 450 | |
| | high | 450 | 440 | 460 | |
| | highest | 450 | 420 | 470 | 0.275 |
| Sodium, mmol/L | | | | | |
| | lowest | 140 | 130 | 140 | 0.908 |
| | low | 140 | 130 | 140 | |
| | high | 140 | 130 | 140 | |
| | highest | 140 | 130 | 140 | 0.454 |
| Potassium, mmol/L | | | | | |
| | lowest | 3.8 | 3.5 | 4 | 0.931 |
| | low | 3.7 | 3.3 | 3.9 | |
| | high | 3.7 | 3.4 | 4 | |
| | highest | 3.9 | 3.7 | 4.2 | 0.006 |
| Carbon Dioxide, mmol/L | | | | | |
| | lowest | 24 | 22 | 25 | < 0.001 |
| | low | 22 | 21 | 24 | |
| | high | 23 | 21 | 24 | |
| | highest | 22 | 19 | 24 | 0.043 |
| BUN, mg/dL | | | | | |
| | lowest | 12 | 8.3 | 15 | < 0.001 |
| | low | 13 | 8.5 | 17 | |
| | high | 15 | 11 | 26 | |
| | highest | 28 | 19 | 40 | < 0.001 |
| Creatinine, mg/dL | | | | | |
| | lowest | 0.77 | 0.63 | 1 | < 0.001 |
| | low | 0.79 | 0.65 | 0.99 | |
| | high | 1 | 0.79 | 1.4 | |
| | highest | 1.4 | 1.1 | 2 | < 0.001 |
| Anion Gap, mmol/L | | | | | |
| | lowest | 11 | 9 | 12 | < 0.001 |
| | low | 12 | 11 | 14 | |
| | high | 12 | 10 | 13 | |
| | highest | 13 | 12 | 16 | < 0.001 |
| WBC Count, 109 cells/L | | | | | |
| | lowest | 5.6 | 4.1 | 6.6 | < 0.001 |
| | low | 7.3 | 5.9 | 10 | |
| | high | 7 | 5.8 | 8.9 | |
| | highest | 8.7 | 6.8 | 12 | < 0.001 |

| | | | | | |
|---|---|---|---|---|---|
| Hemoglobin, g/dL | | | | | |
| | lowest | 15 | 14 | 16 | 0.059 |
| | low | 14 | 13 | 16 | |
| | high | 14 | 13 | 15 | |
| | highest | 15 | 14 | 16 | 0.314 |
| Hematocrit, % | | | | | |
| | lowest | 44 | 42 | 47 | 0.088 |
| | low | 42 | 38 | 46 | |
| | high | 43 | 39 | 47 | |
| | highest | 45 | 41 | 49 | 0.087 |
| Platelet Count, 109 cells/L | | | | | |
| | lowest | 190 | 160 | 230 | 0.015 |
| | low | 230 | 180 | 280 | |
| | high | 230 | 160 | 260 | |
| | highest | 180 | 140 | 290 | 0.224 |
| LDH, U/L | | | | | |
| | lowest | 310 | 250 | 350 | 0.008 |
| | low | 290 | 230 | 410 | |
| | high | 310 | 270 | 440 | |
| | highest | 460 | 330 | 630 | < 0.001 |
| D-Dimer ng/mL | | | | | |
| | lowest | 630 | 390 | 910 | < 0.001 |
| | low | 830 | 580 | 1400 | |
| | high | 910 | 480 | 1600 | |
| | highest | 1400 | 940 | 3400 | < 0.001 |
| CRP, mg/L | | | | | |
| | lowest | 48 | 23 | 71 | < 0.001 |
| | low | 100 | 61 | 170 | |
| | high | 93 | 56 | 150 | |
| | highest | 150 | 100 | 260 | < 0.001 |
| Ferritin, ng/mL | | | | | |
| | lowest | 350 | 170 | 570 | 0.277 |
| | low | 260 | 130 | 590 | |
| | high | 360 | 210 | 560 | |
| | highest | 850 | 350 | 1400 | < 0.001 |

#### Shapley Value Results

Figures E18 through E20 show Shapley Values for each patient and attribute for each individual classifier that make up the three validated hierarchical tests (predicting ICU admission, ARDS and intubation, respectively). Shapley Values that are negative correspond to a classification of higher risk (higher risk classification corresponds to -1 and lower risk classification corresponds to +1), and larger magnitude implies a greater share of contribution from that attribute to classification. Patients are colored by the risk classification of each classifier and the hierarchical structure that gives the final risk label for each test is shown. A reference line is drawn at ±1/(number of attributes), which corresponds to the Shapley Values (SVs) obtained when all features contribute equally to classification.

The tree component of the first split (initial binary) classifiers was treated separately from the Diagnostic Cortex® component. As described in the methods supplement, the Diagnostic Cortex lends itself quite well to a Monte Carlo sampling method for evaluating SVs. The trees did not fit into this schema. However, these trees only use 5 attributes: the output classification label from the Diagnostic Cortex, race, age, gender, and weight. This small number of attributes allows for exact computation of SVs by retraining the trees for all possible subsets of included features. The result is a SV for each of the five attributes, including the Diagnostic Cortex output label. A large magnitude SV for this label is interpretable as a large contribution from the attributes that were used in the Diagnostic Cortex training to the assignment of risk for that classifier. The SVs for the first Diagnostic Cortex models are given in parallel to those of the trees.

**Figure E18: Violin Plot Flow Diagram for the Shapley Values for the Test Predicting Risk of Developing ARDS**

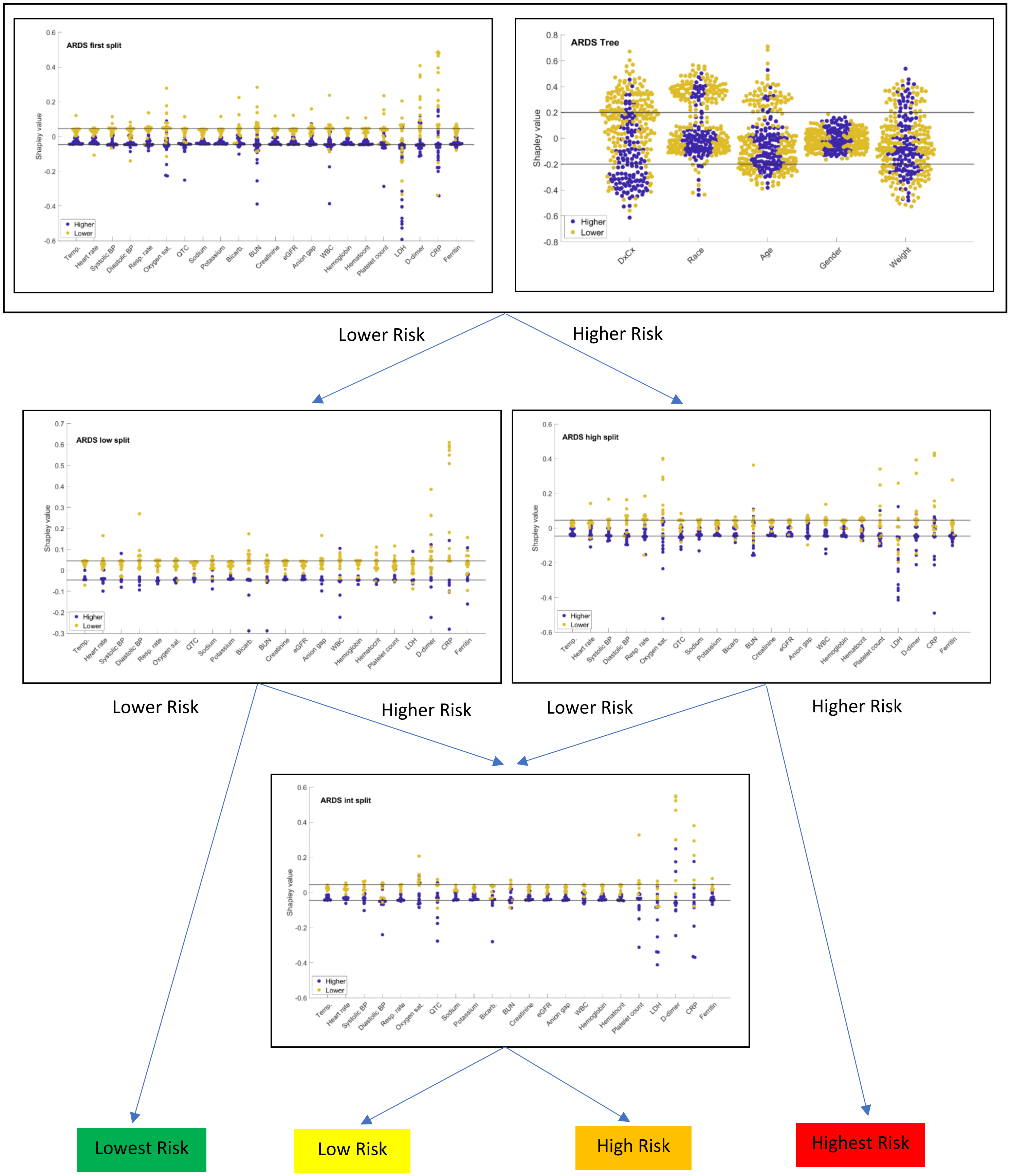

**Figure E19: Violin Plot Flow Diagram for the Shapley Values for the Test Predicting Risk of Admission to the ICU**

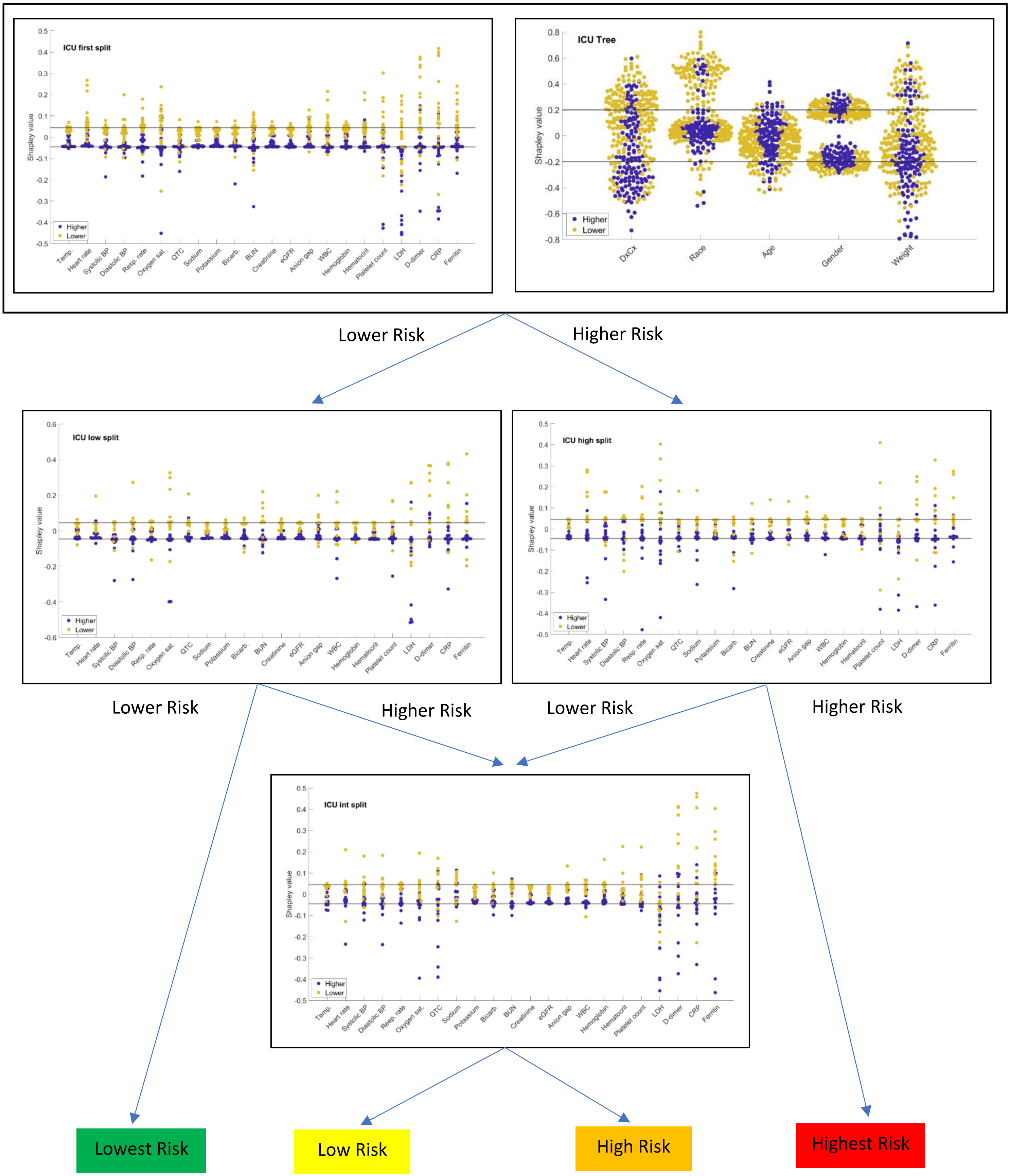

**Figure E20: Violin Plot Flow Diagram for the Shapley Values for the Test Predicting Risk of Intubation**

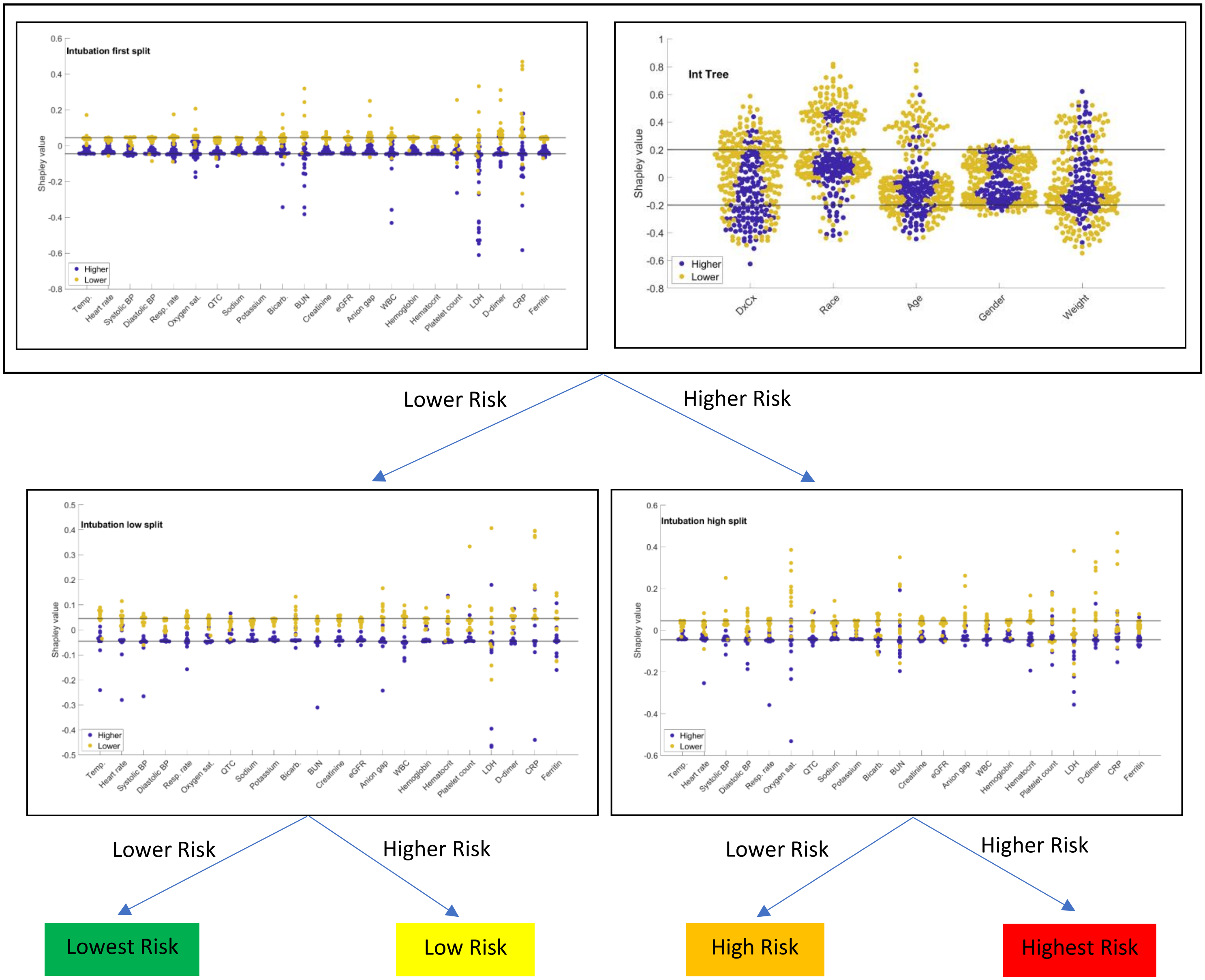

In reading figure E18 showing the Shapley values for the test predicting risk of developing ARDS, some broad conclusions can be drawn. For example, we see that the Diagnostic Cortex attribute in the trees plays a significant role in assigning both higher and lower risk classifications in the initial binary classifier. Gender does not seem to play a role at all, and race, age, and weight play a significant role for some patients. Looking at corresponding Diagnostic Cortex SVs, we see that LDH, CRP, and BUN all frequently contribute to a higher risk classification while CRP and D-dimer frequently contribute to a lower risk classification, in addition to several other attributes to a lesser extent. For patients that are classified as low risk by the initial binary classifier, the low risk child classifier diagram shows CRP and D-dimer contributing significantly again to a lower risk classification. From this, we could conclude that for many patients that get a lowest risk final classification, CRP and D-dimer contributed substantially, with race and age also contributing a fair amount for some patients. Focusing on the highest risk group, the high risk child classifier shows LDH and oxygen saturation contributing to high risk classification of many patients. So, for a final test label of highest risk, LDH, CRP, BUN, and oxygen saturation substantially contribute.

SVs can also be plotted simultaneously with attribute values. Figure E21 shows a scatter plot of SV and CRP value colored by corresponding classification (blue is lower risk and red is higher risk). We see that generally lower values of CRP correspond to lower risk of developing ARDS.

While race/ethnicity was not observed to be among the strongest contributors to risk classification, it was still of interest to investigate further. Figure E22 shows the SVs for race/ethnicity in the first split classifier of the ARDS test separated by race/ethnicity choice. We see that a choice of white corresponds to a contribution towards a lower risk classification while all other choices do not generally contribute towards risk classification.

**Figure E21: Shapley Value vs CRP Value in the ARDS Test First Split**

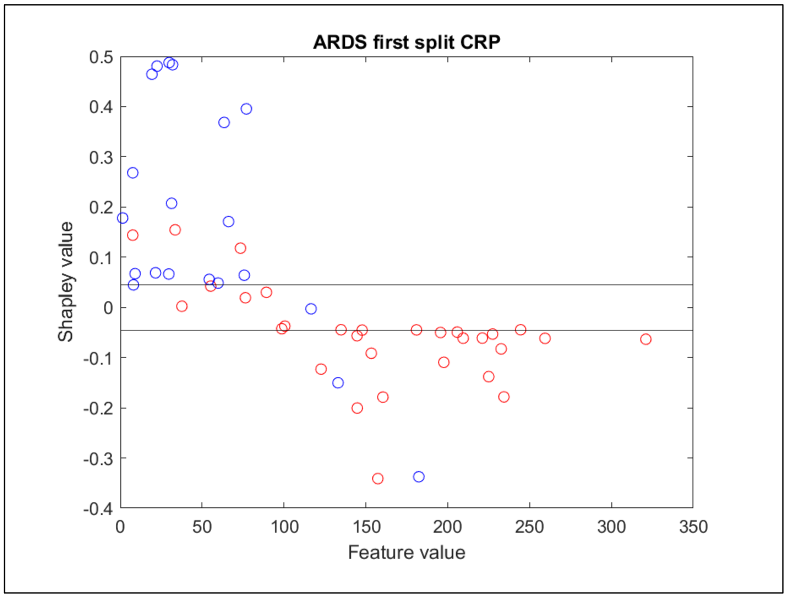

**Figure E22: Shapley Values for Race/Ethnicity by Choice**

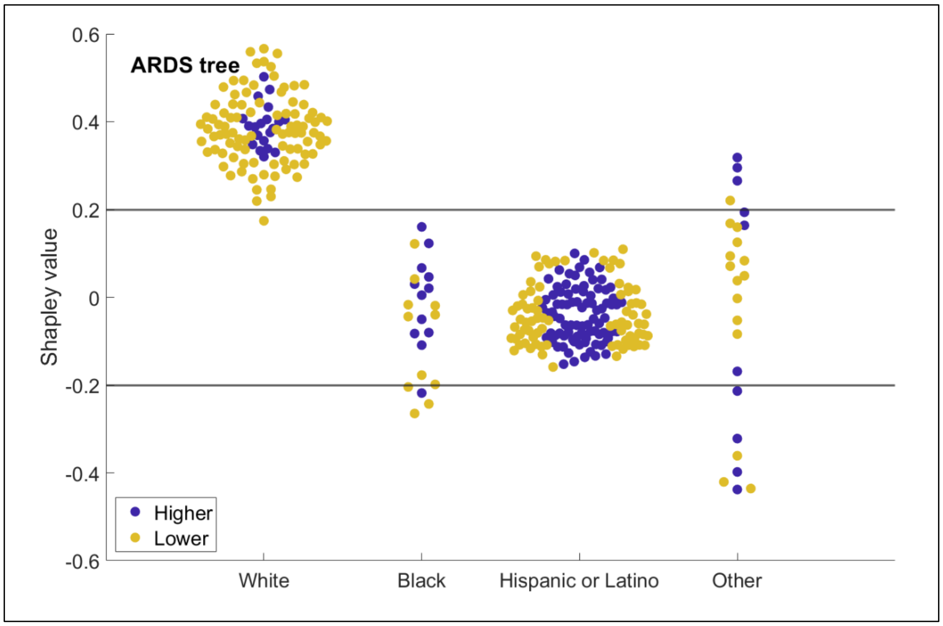


SVs for the test predicting risk of admission to the ICU are shown in figure E19. Again, the Diagnostic Cortex component contributes heavily for assigning both lower and higher risk with weight contributing to a higher risk classification for some patients and race contributing to a lower risk classification for some patients. Gender plays more of a role but is still not as pronounced as the other attributes. The Diagnostic Cortex component of the initial binary classifier shows LDH and CRP playing the largest role in assigning a higher risk classification, but several other features contribute as well. Many attributes contribute to a lower risk classification with ferritin, CRP, D-dimer, LDH, and platelet count seemingly playing more of a role than heart rate, respiratory rate, oxygen saturation and WBC. The low risk child classifier shows CRP and D-dimer contributing to a lower risk classification, so for a final risk label of lowest risk, CRP and D-dimer are perhaps the most important, and ferritin, LDH, platelet count, heart rate, respiratory rate, oxygen saturation, and WBC also contribute. The high risk child classifier shows oxygen saturation, respiratory rate, LDH, CRP, and ferritin contributing to a higher risk classification, so LDH and D-dimer are perhaps the most important with oxygen saturation, respiratory rate, CRP, ferritin, and weight also contributing.

SVs for the test predicting the risk of intubation are shown in figure E20. Again, the Diagnostic Cortex component contributes heavily for assigning both lower and higher risk, with race and age also contributing to a lower risk classification for some patients. The Diagnostic Cortex SVs show LDH, CRP and BUN contributing to a higher risk classification and CRP, D-dimer, LDH, and BUN contributing to a lower risk classification. The low risk child classifier shows CRP contributing to a lower risk classification; so, for a final classification of lowest risk, CRP is likely the most important, with D-dimer, LDH, BUN, race, and age also contributing. The high risk child classifier shows oxygen saturation and LDH contributing to a higher risk classification, so LDH seems to be the most important contributor for a classification of highest risk with oxygen saturation, CRP, and BUN also contributing.

Overall, we see that each of the tests is producing a classification based on a complex interplay of multiple attributes and that, while we can discern some general trends across the cohort, the relative importance of the attributes to classification is dependent on the patient.

The preceding arguments tried to consider how the attributes affected the classification of samples in aggregate. These arguments were fairly general; we believe the information at the patient level is more directly interpretable and can be very informative for personalized treatment options. Figures E23 through E25 show three examples of how results might be presented for an individual prediction using the test predicting risk of intubation. Interpretations are given in the comments. In each of the individual diagrams, the SVs are given with a reference line at 1/(number of features). Color indicates higher (red) or lower (green) risk in the case of the initial binary (first split) classifier and final classification level: highest, high, low, or lowest (red, orange, yellow, or green) for the child classifiers.

**Figure E23: Individual SVs for patient ID 105149887**

Patient ID: 105149887

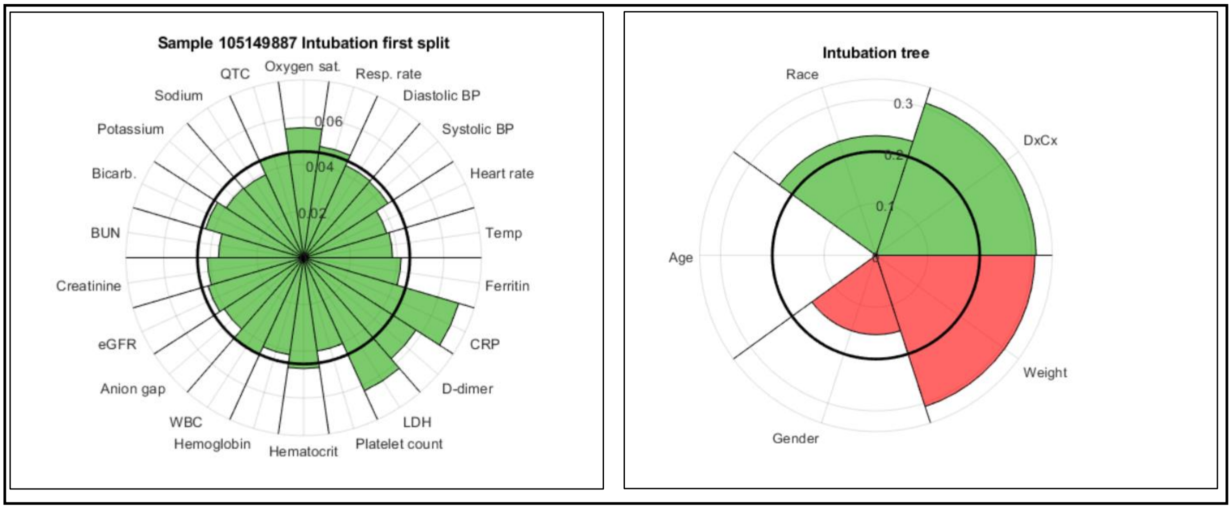


**Final Classification: Low Risk (yellow)**

**Comments:**

Patient at lower risk of intubation in first split from CRP, D-dimer, LDH, and oxygen saturation, but higher risk from weight. Child low risk split shows higher risk from BUN anion gap, respiratory rate, BPs, heart rate, and temperature, but lower risk from hematocrit, CRP, and LDH.

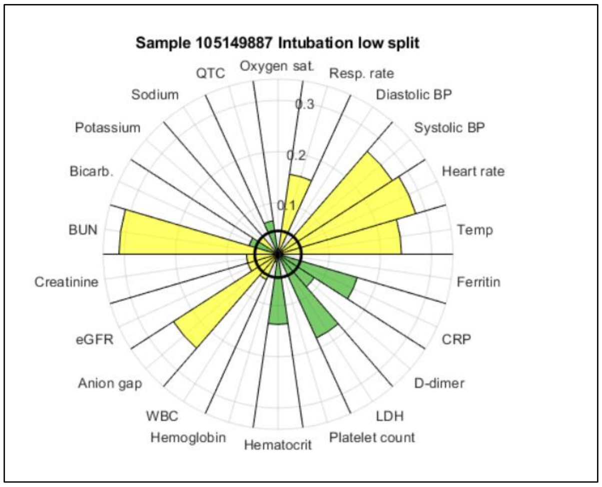

**Figure E24: Individual SVs for patient ID 19675902**

Patient ID: 19675902

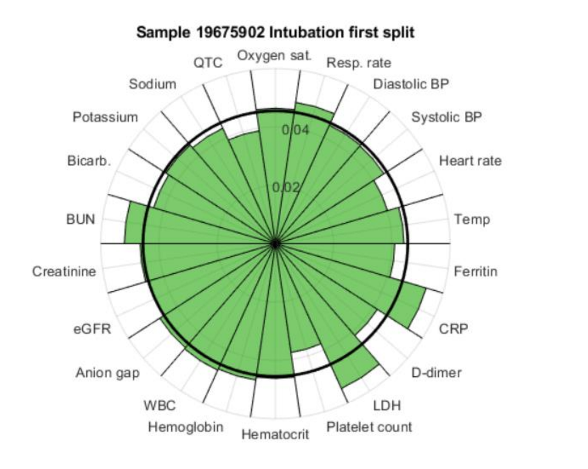


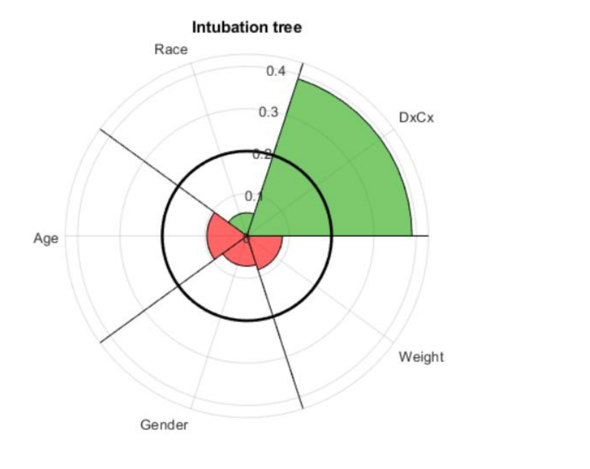


**Final Classification: Lowest Risk (green)**

**Comments:**

Patient has no attributes signaling higher risk of intubation and no attributes strongly signaling lower risk compared to all the others.

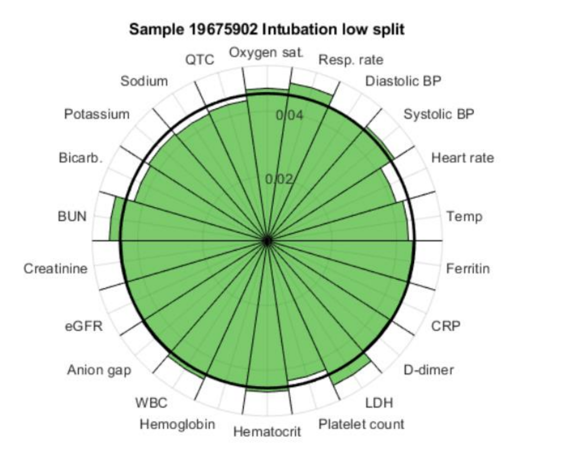

**Figure E25: Individual SVs for patient ID 331603541**

Patient ID: 331603541

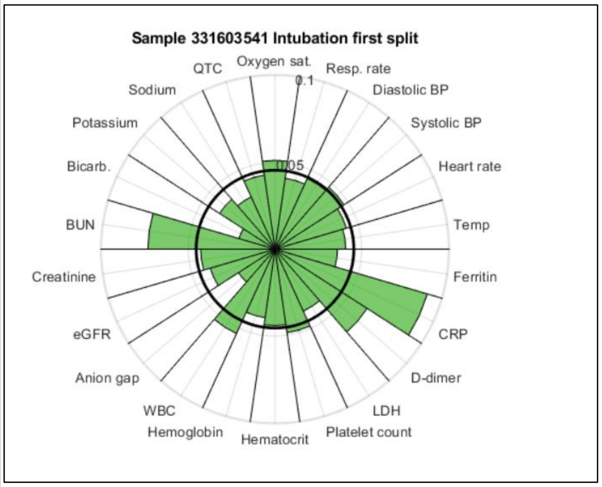


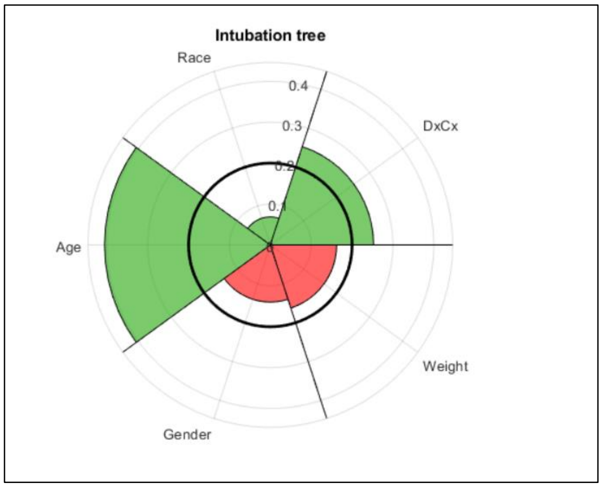


**Final Classification: Lowest Risk (green)**

**Comments:**

Patient at lower risk of intubation due to age, CRP, and BUN at first split and lower risk at second split due to CRP and ferritin.

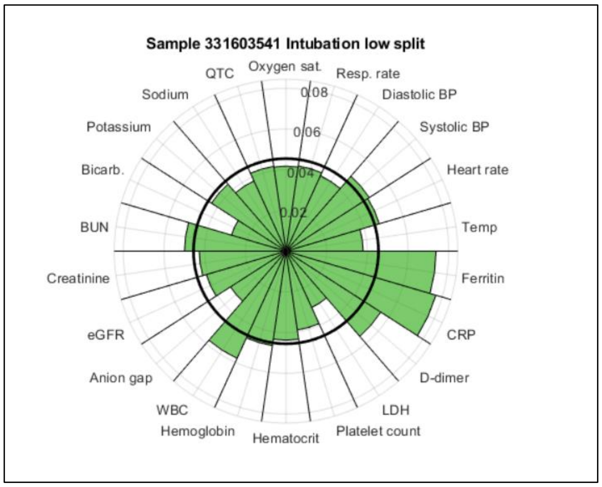


References

[E12] Cohen J. Statistical Power Analysis for the Social Sciences (2nd Edition). Hillsdade, NJ, Lawrence Erlbaum Associates; 1988.

[E13] Breiman L. Out-of-bag estimation: Technical Report. Department of Statistics, University of California; 1996.
